# Exploring global and specific pathogenic mechanisms in Chronic Chagas Cardiomyopathy through multi-omics integration

**DOI:** 10.1101/2023.10.23.23297068

**Authors:** Pauline Brochet, Jorge Kalil, Vincent Procaccio, Edecio Cunha-Neto, Lionel Spinelli, Christophe Chevillard

## Abstract

Chagas disease is a neglected disease from South America caused by a parasite, *Trypanosoma cruzi*. While most of infected people remains asymptomatic, around 30% develop Chronic Chagas Cardiomyopathy (CCC), a very lethal cardiomyopathy characterized by an exacerbate inflammatory response. The last few years, our team has set up multiple omics analysis. Briefly, we have pointed the over-expression of many genes involved in the Th1 lymphocyte response, as well as some epigenetic features potentially involved in their regulation, including miRNA, lncRNA and methylation site. Moreover, some mitochondria mutation seems to predispose to the development of CCC. In order to understand and characterize the impact of genetic and epigenetic elements on the pathogenic process associated to CCC, we have performed here a multi-omics integration, combining transcriptomic, methylomic, miRNomic and mitochondria sequencing. We have identified two distinct pathogenic pathways that vary among patients with chronic Chagas cardiomyopathy (CCC). One pathway is primarily influenced by IRF4, a transcription factor known for its involvement in the development of both B and T cells, while the other is driven by TLR signaling. Notably, genes related to B cells play a role in both of these processes. Additionally, we have detected certain similarities in the B cell receptors of all CCC patients, which may potentially contribute to autoimmunity. While further analysis is necessary to validate these findings, they collectively enhance our understanding of the pathogenic mechanisms associated with CCC.

## Introduction

Chagas disease is a parasitical disease caused *Trypanosoma cruzi*, a parasite endemic from South America, and spreading across the globe with the migratory flow (1–3). After the infection, the acute phase is mostly asymptomatic, and most of the contaminated peoples remains asymptomatic (ASY) through the years. Nonetheless, approximately 30% develop Chronic Chagas Cardiomyopathy (CCC), the main cause of death of Chagas disease, which kill 10.000 peoples each year. At this stage, the parasitemia is very low, and anti-parasite treatments have no impact on heart deterioration (4).

CCC is characterized by a myocardial dilatation and an infiltrate of immune cells (mainly lymphocytes and macrophages), inducing continuous inflammation (5). Although the mechanisms driving the development of this cardiomyopathy remains unclear, there is a notable disparity in the differentiation of Th lymphocytes into Th1 and Th2 cells in CCC, which results in a prevalent Th1 profile in CCC patients and a high production of pro-inflammatory cytokines, including IFN-γ and TNF-α. Conversely, asymptomatic (ASY) patients present a more equilibrium lymphocytes Th profiles, with a higher production of anti-inflammatory cytokines like IL-10 (6). Furthermore, mitochondrial dysfunctions have been detected in CCC, leading to reduced mitochondrial activity and the consequent production of Reactive Oxygen Species (ROS). While during the acute phase, ROS serve to eliminate parasites, their prolonged accumulation in cardiac tissue eventually leads to tissue damage (7). The hypothesis of an autoimmune reaction developing in CCC patients has been proposed, supported by the identification of auto-reactive peptides that stimulate B or T cells in human sera (8).

With advances in large-scale analysis techniques, many study have focused on understanding the pathogenic process leading to the development of CCC (9). To this end, our team has carried out genomic, transcriptomic and methylomic studies highlighting processes specific to CCC patients. Focusing on mitochondrial DNA, only a limited number of mutations linked to cardiac diseases have been identified. However, there is evidence that certain mitochondrial haplogroups are overrepresented among CCC patients. Transcriptomic studies have revealed the involvement of immune-related genes, including Toll-like receptors, Th-1 lymphocyte genes, and cytokine signaling, when compared to healthy asymptomatic, dilated cardiomyopathy (DCM) controls (10,11). Interestingly, some genes associated with cardiac and nervous system processes also exhibit dysregulation between CCC and DCM, illustrating the specificity of this disease. Some associations have been established between the expression of these genes and epigenetic regulatory elements, including non-coding genes such as miRNA (12) and lncRNA (13), and methylation sites (11,14). While identified ncRNA are mostly involved in heart-related processes (miR-1, miR-133a, miR-208b, MIAT), DNA methylation mainly occurs in genes involved in immune response, nervous system and ion transport. The binding of some transcription factors, including PAX5, RUNX3, SPI1, IRF4 and TBX21, was predicted to be altered by a differential methylation in targets promoter region, supporting the impact of methylation in this disease.

Although these studies offer an initial analysis of the factors contributing to the development of CCC, they are constrained to the combination of only two types of omics data, providing only a partial perspective on the underlying mechanisms. To get a more complete comprehension of the pathogenic process occurring in CCC myocardium, we performed here a multi-omics integration of transcriptomic, miRnomic, methylomic and mitochondria sequencing data.

## Materials and methods

### Raw data collection

All raw data comes from our previous studies, including bulk RNA-sequencing (11), miRNA sequencing (12), Illumina EPIC methylation (11) and mitochondria variant calling. Human left ventricular free wall heart tissue samples were obtained from patients with end-stage heart failure CCC at the time of heart transplantation (n=7). CCC patients underwent a serological diagnosis of *T.cruzi* infection and standard electrocardiography and echocardiography, and tissues were subject to histopathological assessment as previously described. The protocol was approved by the institutional review boards of the University of São Paulo School of Medicine and INSERM (French National Institute of Health and Medical Research). Written informed consent was obtained from all patients. All experimental methods comply with the Helsinki Declaration. Each dataset was aligned on the GRCh37 (hg19) human reference genome from Ensembl, and all classical pre-processing steps have been applied as described in previous study (11,12).

### Missing samples imputation

Across the multi omic dataset, 6 samples are missing in at least one omic: 4 in transcriptomic and 3 in miRNomic (**table 1**). To complete the multi-omics dataset, missing values for samples were imputed using the KNN-imputer function of scikit-learn python library. All features’ values were centered and scaled to be comparable, and a custom matrix distance for the multi-omics dataset was made, considering for each omic: the distance between all samples, the number of features and the number of missing samples. The imputation of controls and cases was done independently. For each phenotype, the following steps have been performed:

*Step 1.* For each omic M and for each couple of samples (i,j) in M, Euclidean distance metrics has been computed:

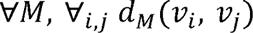

*Step 2.* For each distance matrix obtained from step 1, each distance value between a couple of samples (i,j) was divided by the number of features N_M_ in omic M. Since the number of features is very different from one omic to another, this step allows to normalize the distance between two samples in each omic by the number of features:

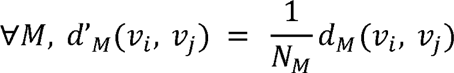

*Step 3.* An average distance matrix has been computed. The normalization term of the mean corresponds to the number of available samples through omics:

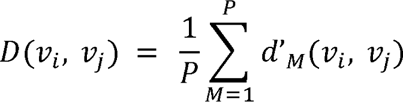

*Step 4.* The missing samples imputation is performed with the KNN imputer function of sklearn python 3 package (v.1.2.2), using the custom distance matrix made previously.

By this way, both differences in omic size and number of missing samples are normalized in the final distance matrix, and all omics have the same contribution in data imputation.

### Data augmentation

An estimation of the optimal number of samples to ensure a statistical power greater than 0.8 for the multi-omics integration was performed using MultiPower (v.1.0). To reach the desired numbers, synthetic samples were created as linear combinations of randomly selected original samples of the same phenotype. For each synthetic sample, random weight between 0 and 1 were drawn for each selected sample so the sum of all weight is reach 1. To ensure synthetic sample remain in a region of the samples space that can have a biological meaning the first weight, called α, was chosen to be between 0.8 and 1. If we note {P_i}_i=1,..,K_ a sampling of K samples in the N original samples and P’ the new synthetic sample we have:

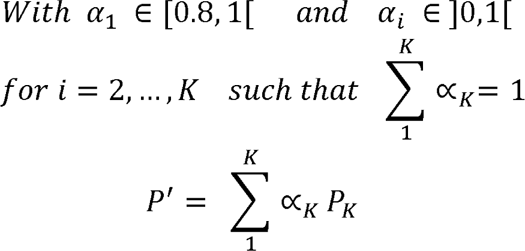

In total, 193 synthetic samples were generated per phenotype, to obtains a total cohort of 400 samples.

### Feature reduction

Given the large number of features in some omics, a feature selection was applied to both tanscriptomic and methylomic datasets. For the transcriptomic features, a filtering process was applied based on gene function, retaining only transcription factors, protein-coding genes, and long non-coding RNA (lncRNA). A second feature selection was performed in lncRNA, keeping only those annotated in lncRNAdisease (15). In the case of methylation sites (CpG), due to their large quantity, only CpGs located in regulatory regions of genes (TSS-1500 and 5’UTR region) included in ChagasDB (9) were included in the analysis.

### Multi-omic integration

The multi-omic integration was conducted using MOFA+ (v.1.8.0) (16) with the selected features. All the following analysis was performed using R (v.4.2.1). For transcriptomic expression data (RNA sequencing and miRNA sequencing), a log2 transformation was applied to ensure a Gaussian distribution. CpG methylation beta values were transformed into M-values (17) and modeled using a Gaussian likelihood, similar to the approach employed in the original MOFA+ analysis (16). The variant allele frequency of mitochondria was binarized, representing the presence or absence of the mutation, and modeled using a Bernoulli distribution. The analysis was iterated 19 times, gradually increasing the number of factors from 2 to 20. To ensure the selection of meaningful factors, only those representing at least 1% of the data variability within a single omic were retained. The specific function of each factor was evaluated using MOFA GSEA enrichment on Reactome database (18). For each factor of interest, the weights of features within that factor were scaled individually by omic. Only the top features with sum of weights equal to or greater than 25% of the total feature weights within that specific factor were retained. A similar analysis was performed using CCC patients only. In this case, the features weight cut-off was fixed to 50% of the total feature weights.

### Pre-processing pipeline validation

The quality of the data augmentation was tested using 1643 samples from TCGA (19), including breast (BRCA n = 1106) and lung cancer (LUAD, n = 537) transcriptomic data (**supplementary table 1**). For each dataset, different oversampling methods were applied, using each time 10 existing samples to generate 200 synthetic samples. The other methods include bootstrapping, SMOTE (v.0.4.1) (20) and KNNOR (v.0.0.4) (21). The validity of the results obtained with MOFA was assessed by comparing them with those obtained with 3 other multi-omics integration methods: DIABLO (MixOmics) (v.6.24.0) (22), JIVE (v.2.4) (23) and MCIA (v.1.40.0) (24). Using the same filters as before, the obtained features were compared using the Jaccard index, and the overlap coefficient. Finally, the complete analysis was performed 10 times. For each run, one raw sample was randomly removed in one omics. The results were compared using the Jaccard index, and the minimal overlap.

### Multi-omics correlation network

In order to understand the link between the selected multi-omics features, two correlations networks were made, specific for each phenotype (healthy controls and CCC). The transcriptomic data was split into 3 different types of features: protein coding genes, transcription factor and lncRNA. Correlation clusters were assessed using the pheatmap R package (v.1.0.12) distinct biological functions associated with each cluster were identified using the R gprofiler2 package (v.0.2.1). To ensure the reliability of the identified correlation, the network were filtered using interaction public databases, including STRING (25), BioGRID (26), Coexpedia (27), GRNdb (28), IntAct (29), lncTarD (30), miRecords (31), miRTarBase (32) and TarBase (33).

### Lymphocyte T and B repertoire analysis

An analysis of immune repertoire was performed with TRUST4 (v.1.0.8) tool on tissue RNA-seq samples (7 control, 10 severe CCC and 8 dilated cardiomyopathy) (34). TRUST4 align reads against V, D, J and C genes, and exports all the distinct clonotypes found in each sample. These clonotypes were then analyzed thanks to the ImmuneArch R package (v.0.8.0), in order to identify shared clonotypes and gene usages between samples of the same phenotype (35). Clonotypes sequences were clustering using Clustal Omega (v.1.2.4) (36).

## Results

### Validation of the pre-processing pipeline

Multi-omics analysis was conducted on a total of 14 human heart tissue samples, comprising 7 healthy controls and 7 CCC cases, and 4 different omics (transcriptomic, methylomic, miRnomic and mitochondria sequencing). Based on multi-power tool estimation, the statistical power using so few samples fall within the range of 0.1 to 0.3, depending on the specific omics under evaluation. According to this estimation, it was necessary to increase the sample size to a minimum of 75 samples per phenotype to achieve a minimal statistical power of 0.8 in each omic (**supplementary figure 1A**). Therefore, synthetic samples were generated using the barycenter metrics, as detailed in the materials and methods section. To confirm the stability of the analysis result, synthetic datasets of increasing sizes (from 21 to 400 samples per phenotype) were created and submitted to a multi-omics integration. When the select features after multi-omics integration with the largest cohort (n = 400) were compared with varying lower sample counts, it was observed that the same features were consistently found for 100 samples and more (**supplementary figure 1B**). Given the high number of features in different omics, we opted to double the sample size to 200 per phenotype, aiming to achieve a statistical power of almost 1 for each omic, as a statistical power of 0.8 might prove insufficient. Compared to other existing methods, the barycenter approach better preserves the raw data distribution observed in PCA compared to bootstrapping and KNNOR and is similar to SMOTE (**supplementary figures 2**). About the multi-omics integration itself, it has been performed using the MOFA+ methods, which provides similar results to JIVE and MCIA (**supplementary figure 3A**), with 40% (1361/3338) of MOFA+ features in common with JIVE or MCIA. Particularly, 70% of MOFA+ features were retrieved in MixOmics integration, 96% in JIVE and 41% in MCIA (**supplementary figure 3B**). Those overlap, considering the vast number of original features, strongly indicate that the selected features were not randomly chosen and hold relevance for studying CCC. To validate the effectiveness of the overall pipeline, we repeated the process 10 times using different generated datasets, each time excluding one existing original sample. By this way, the values of this sample will be imputed, and some synthetic samples will be generated from his imputed values. All iterations produced consistent outcomes, with a minimum Jaccard index of 70% and a minimum overlap coefficient of 90% in shared features (**supplementary figure 4**). These consistent results strongly support the robustness of this pipeline (**figure 1**).

**Figure 1.**
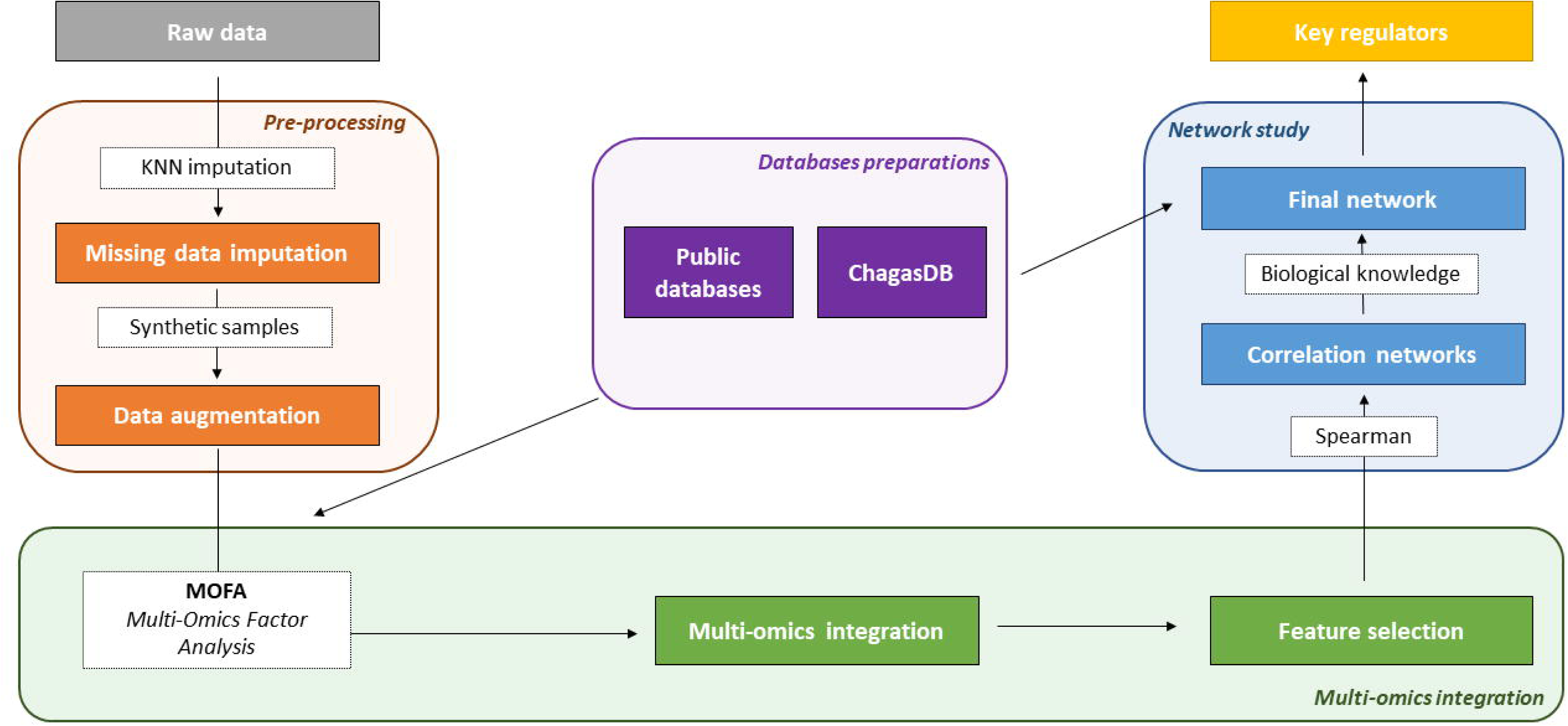
Workflow overview. Raw data were obtained on end-stage CCC heart tissue and healthy heart tissue. All the individual omics pre-processing, from alignment to normalization, were performed as described in previous study. Multi-omics pre-processing steps (orange) include missing samples imputation with KNN-imputer from scikit-learn library, and synthetic samples generation using barycenter approach. The multi-omics integration itself was performed using MOFA+ tool (green). The features selected at multi-omics level were used to create phenotype-specific correlations network (blue). The downstream analysis was made on correlation network filtered with known or predicted interactions in public databases.

### Factor 1 is associated with Chagas disease Cardiomyopathy

The MOFA integration was conducted on a subset of the complete dataset, consisting of 266,256 features across the 4 omics (transcriptomic n = 19,559, methylomic n = 246,660, miRnomic n = 652, mitochondria sequencing n = 45). According to MOFA integration, 7 factors carry at least 1% of the data variability. The 2 first factors carry the most variance in different omics. Whereas factor 1 is strongly supported by transcriptomic and methylomic data, factor 2 is carried by mitochondria sequencing (**supplementary figure 5A**). No correlation was found between pairs of factors, indicating each of them is carrying specific information (**supplementary figure 5B**). Whereas 6 factors display a significant difference between CTRL and CCC weight (Wilcoxon p-value ≤ 0.05), the factor 1 is the most discriminative between CCC and healthy controls (**figure 2A**). Moreover, most of the top-rank genes in factor 1 are known to be involved in Chagas disease cardiomyopathy, which is less the case for other factors (**figure 2B**). Interestingly, some of the top-ranked features are absent in ChagasDB, and therefore, remain unknown in the context of this disease. Considering the interest of factor 1 in this disease, a GSEA enrichment was performed using all the features rank in this factor, using the Reactome database. The positive features, so the features positively enrich in CCC, are involved in both innate and adaptative immunity (Complement cascade, TCR/BCR signaling…) (**supplementary figure 6**). Interestingly, the interleukin-10 signaling, associated to anti-inflammatory response, is also enriched. On the opposite, some cardiac and mitochondrial processes are negatively enriched. All these results are in line with the current knowledge of CCC. Considering the importance of factor 1 on the disease, the top-features associated to this factor were selected, including 1,214 genes, 2,066 methylation site and 58 miRNA. The majority of the selected genes are involved in immune response, mostly associated to T cell and inflammatory response (**supplementary table 2**). However, most of top-ranked positive features are known to be involved in B-cell related process (FCRL5, IGJ, CLNK, FAM30A, NUGGC, CD79A, IGLL5), suggesting an important role of this cell in CCC.

**Figure 2.**
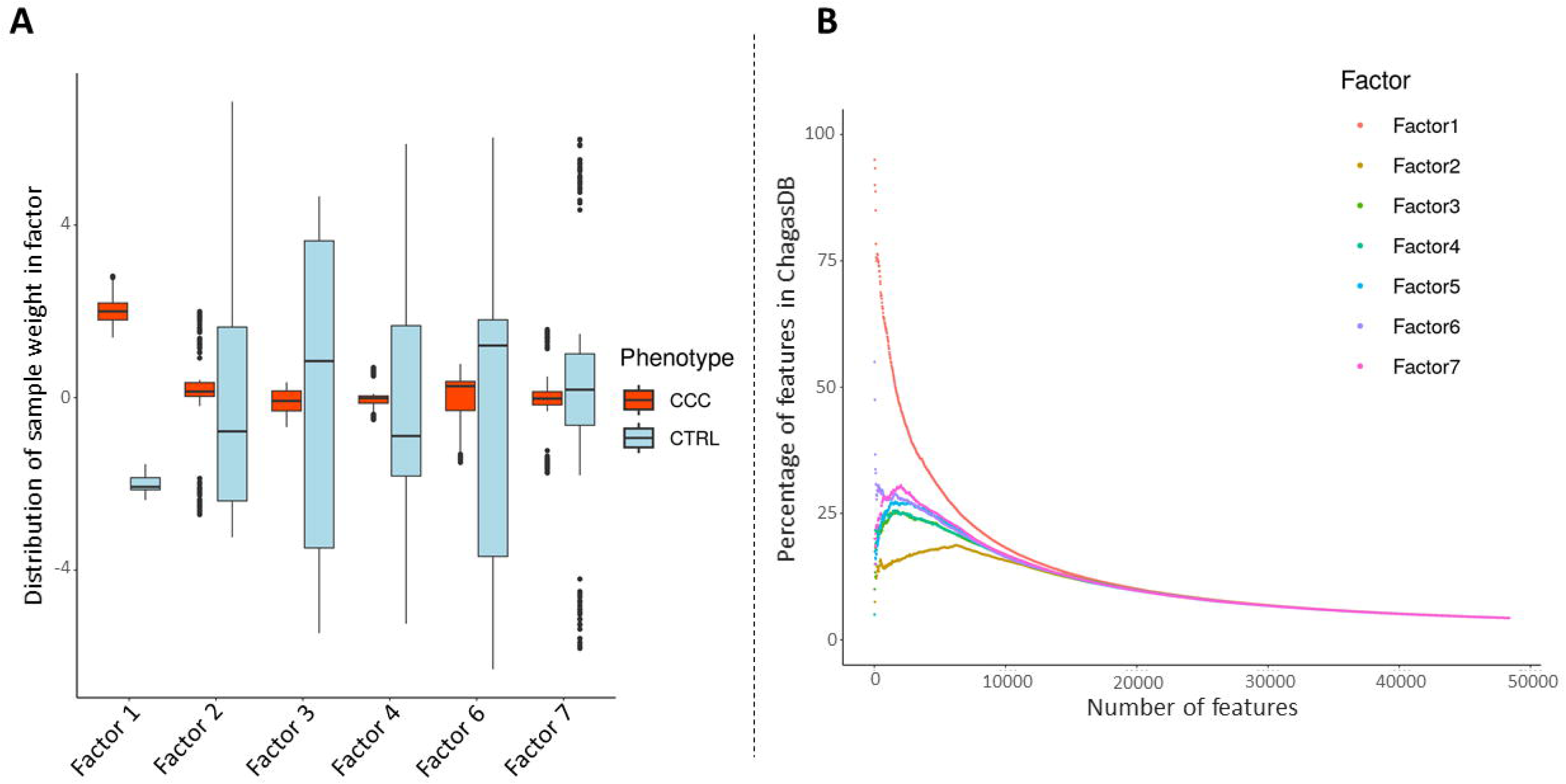
**(A)** Distribution of sample contribution through the 6 factors displaying a significant difference between CTRL and CCC weight in each factor (blue: CTRL, red: CCC). (**B)** Percentage of features present in each factor within ChagasDB, arranged based on their respective weights in each factor.

### CCC patients presents specific regulatory patterns

To identify deregulated processes in CCC, two correlation networks were constructed, one based uniquely on healthy control samples, and the other only on CCC samples. Only correlations between expressed features were retained, with methylation sites assigned to the associated gene. To investigate the potential involvement of regulatory elements, the transcriptomic dataset was divided into three types of features: protein coding, transcription factors (TF), and long non coding RNA (lncRNA). Interestingly, the correlation levels between some pairs varies depending on the phenotype (**supplementary figure 7A**). Given these observations, the correlation networks were filtered as follows: the absolute correlation level must be over 0.8 in one phenotype, and less than 0.4 in the other (**supplementary table 3**, **supplementary table 4**). Although the same features are present in both networks, their behavior is significantly different between CCC and CTRL (hypergeometric p-value <= 2E-16) (**supplementary figure 7B/7C**). Looking at the different impact of regulatory features on gene expression, we perform a hierarchical clustering (HCA) to group them according to their correlation with protein-coding genes. 3 specific clusters appear, illustrating clear different co-expression patterns (**figure 3A**) (**supplementary table 5, supplementary table 6**). This phenomenon is not observed in healthy controls (**figure 3B**). Using KEGG and Reactome database, we identified 47 pathways significantly enriched in at least one cluster (**figure 3C**). Features in cluster 2 are associated to T-cell related processes (T cell receptor signaling pathway, Th1 and Th2 cell differentiation, Th17 cell differentiation), whereas cluster 3 is associated to B cell response (B cell receptor signaling pathway) and innate immune response (Toll-Like Receptor, complement activation). None of the KEGG or Reactome pathways were specific to the cluster 3. Interestingly, pathways related to anti-inflammatory response, including IL-10 and PD-1 signaling affiliated to cluster 2. Moreover, some autoimmune diseases (type 1 diabete, inflammatory bowel disease…) were also associated to cluster 2, as well as viral myocarditis.

**Figure 3.**
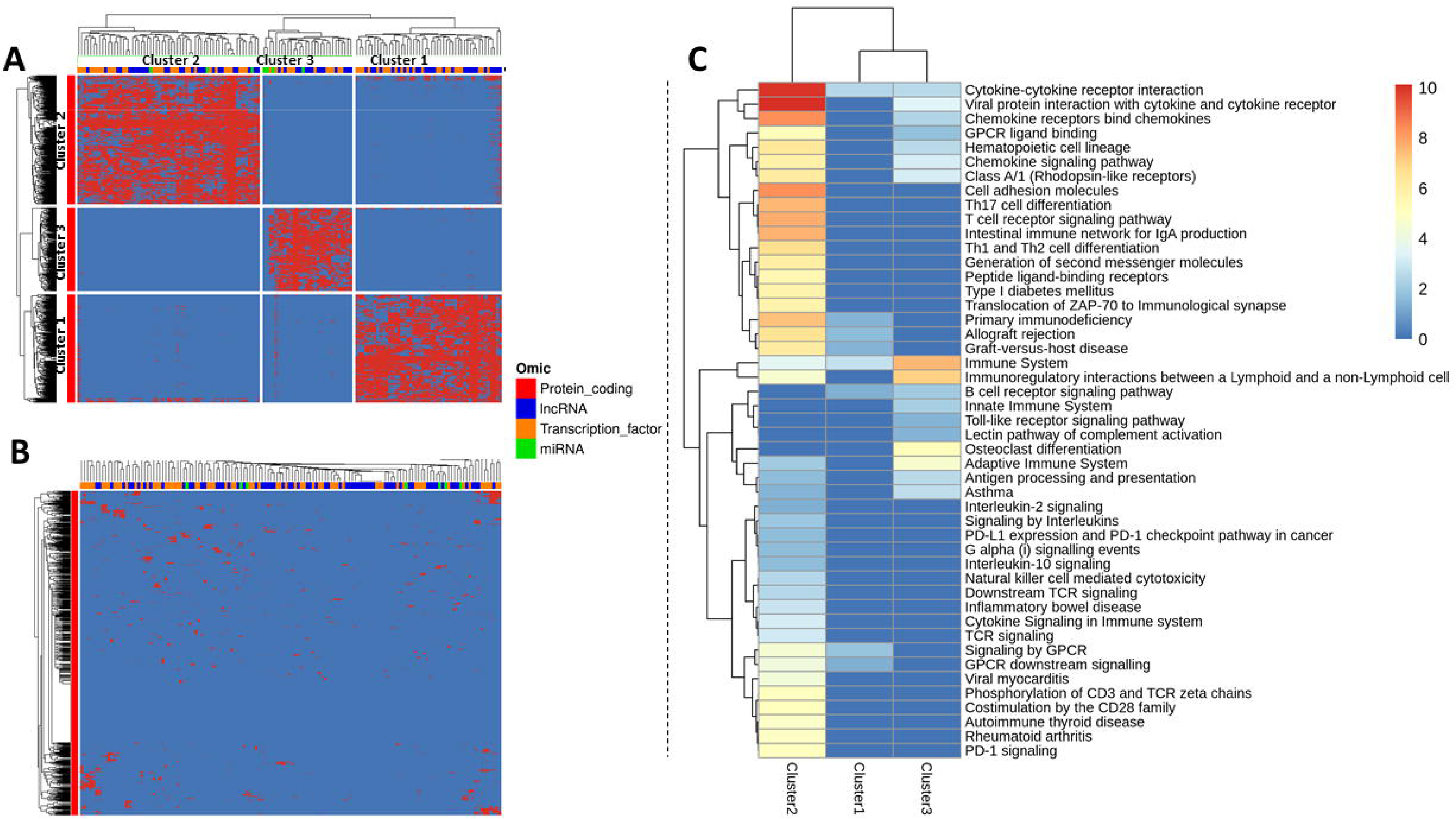
**(A)** Hierarchical clustering of the regulatory features (lncRNA, miRNA, transcription factor) correlated with protein coding genes in CCC network. **(B)** Clustering of the regulatory features (lncRNA, miRNA, transcription factor) correlated with protein coding genes in CTRL network. **(C)** KEGG and Reactome pathway significantly enriched in at least one of the CCC network cluster.

### B cell receptors present a high convergence in all CCC

Considering the previous results highlighting the importance of B and T cell-receptor signaling in CCC, an analysis of BCR and TCR repertoire was performed. The TCR repertoire analysis uncovered that the diversity and abundance of clonotypes were higher in CCC than in DCM or CTRL (**supplementary figure 8A**). However, no clonotype was found to be shared across all CCC (**supplementary figure 8B**).

Analysis of the BCR heavy and light chains revealed that the diversity and abundance of clonotypes was much greater in the CCC samples than in the CTRL or DCM samples (**figure 4A**). This result is a first control that confirms an increased presence and/or activity of B cells in the samples, in association with chronic inflammation. The specific study of light chain clonotypes by considering the CDR3 sequence and the corresponding V/J genes reveals that many clonotypes are common to several CCC samples and that more than 10 of them are common to all 10 samples (**figure 4B**). This phenomenon is not found in the CTRL or DCM samples in which the clonotypes are shared by a maximum of 2 samples. The detection of this public clonotype repertoire seems to indicate the presence of sequence convergence by specific selection, similar to that found in the presence of specific antigens. However, the absence or very low parasitemia in the patients’ cardiac tissue seems to rule out the possibility of the presence of pathogen antigens. In order to be more precise, we wanted to check whether this convergence was found in a larger proportion of clonotypes and whether similar but not identical sequences to the public repertoire could be detected.

**Figure 4.**
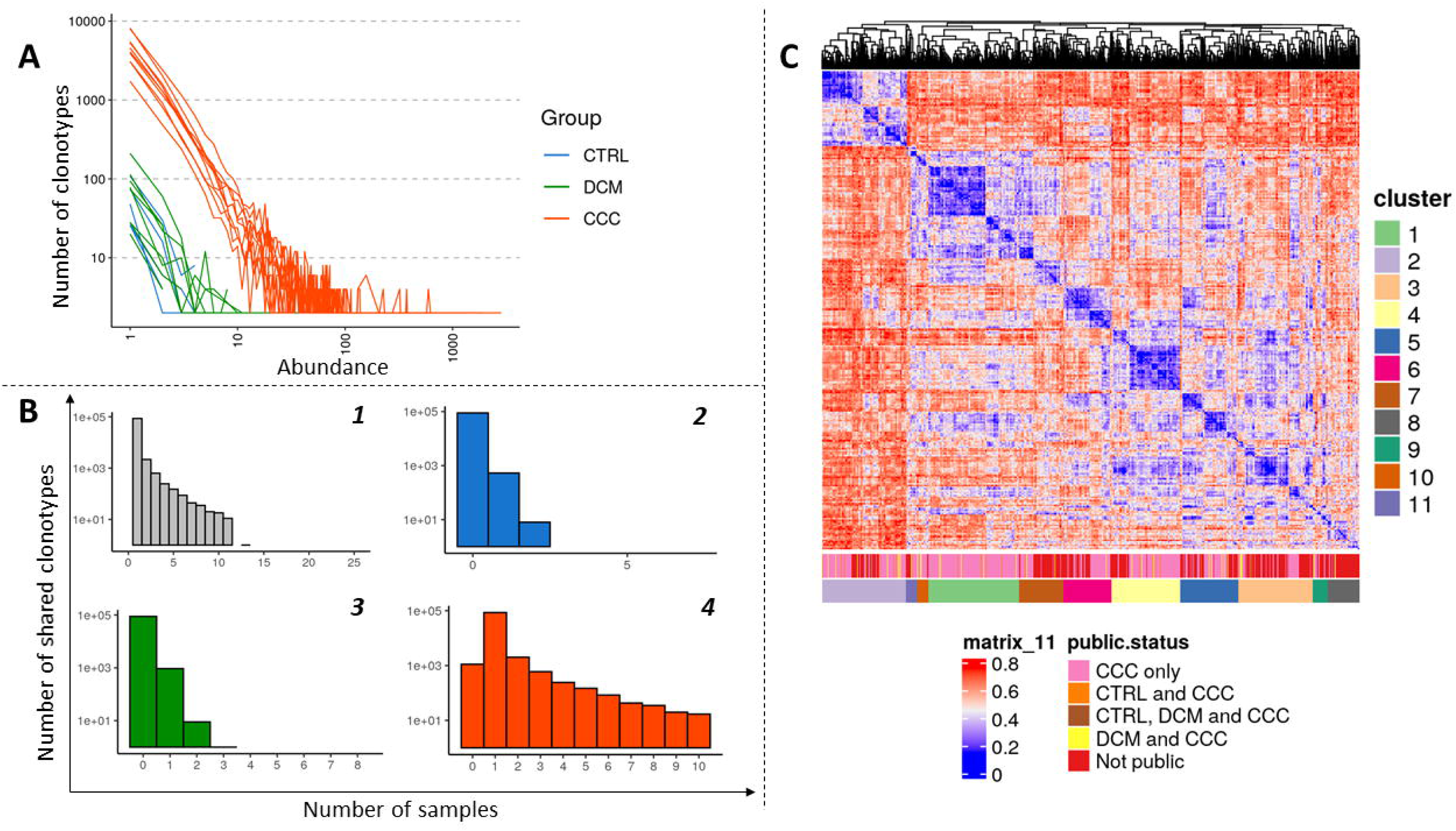
Clonotypes analysis of B cell receptor. **(A)** Abundance of the BCR clonotypes per phenotype. **(B)** Distribution of the number of samples sharing public clonotypes per phenotype: **1)** All samples, **2)** CTRL samples, **3)** DCM samples, **4)** CCC samples. **(C)** Hierarchical clustering of 4000 clonotypes (2000 shared by at least 2 samples and 2000 unique). The phenotype concerned and the resulting clusters are indicated by color legends.

We thus selected 2000 clonotypes shared by at least 2 samples and having the longest CDR3 sequences. As a control, we then selected 2000 clonotypes present in a single sample and having the longest CDR3 sequences. Using Clustal Omega, we aligned the sequences and then calculated the distances between these 4000 sequences (**figure 4C**). The distance matrix clearly shows the convergence of clonotype series around different clusters. We used an HCA algorithm to cluster the sequences by their proximity. 11 clusters were thus identified (with an arbitrary choice of cutting the hierarchical tree). For each cluster, we studied the frequency distribution of the V genes associated with the sequences. The majority of clusters are dominated by the presence of one or two V genes, often close to each other (**supplementary figure 9**). Those clusters are also dominated by clonotypes of the public repertoire. With regard to the BCR heavy chains, while a strong clonal expansion was always observed in the CCC samples but not in the CTRL or DCM samples, the presence of a public repertoire was not observed. The stochiometric relationship between light and heavy chains remains close to the expected value (expected value: 1.5, observed value: 1.17). As the heavy chain is commonly more mutated than the light chain, it is possible that the sequencing depth we used was not sufficient to reveal the commonality of certain sequences, given the very large possible repertoire.

### Dual and opposite role of lymphocytes-related transcription factor

To identify potential regulatory interactions among the correlated features, we compared our correlation networks with various interaction databases (see Methods). Both healthy control and CCC specific correlations are significantly enriched in known interaction (CTRL hypergeometric test p-value p-value < 3.68E-249, CCC hypergeometric test p-value < 2.87E-08), confirming their relevance. Consequently, we filtered the correlation networks, keeping only correlated features known or predicted to interact together. With the aim of characterizing the mechanisms that contribute to the exacerbation of the inflammatory response in CCC, we focused on the correlations/interactions between different type of features previously assigned to cluster 2, mainly involved in T cell activation and TCR signaling. As expected, the most interconnected features are protein coding genes and transcription factors, whereas miRNAs and lncRNAs have a limited number of targets. For example, RP11-357H14-17, the top-correlated lncRNA assigned to cluster 2, was removed from the interaction network. Looking at the 10-top connected TF, all of them are related to T- or B cell activation and development. Moreover, they seem to combine to regulate distinct sets of genes (**supplementary figure 10A**).

We have identified 9 TF that exhibit correlations with TCR genes, suggesting activation through the TCR signaling pathway. Notably, 7 of them also show correlations with TLR and/or BCR genes. Furthermore, they are associated with genes participating in 9 specific biological functions of interest (**figure 5A**). IRF4, IKZF1, and PAX5 are linked to all these processes. IRF4 displays an inverse correlation with certain cardiac-related genes, such as MYH6 and RRAD (**figure 5B**). On the opposite, it exhibits a positive correlation with genes involved in inflammation regulation, as well as other lymphocytes-related TF (TBX21, PRDM1, BATF, PAX5, IKZF1) (**figure 5C**). Among the genes related to this process, RHOH is the only gene that is downregulated in CCC. MIR551b demonstrates an inverse correlation with SOCS3, suggesting a potential involvement of this miRNA in the regulation of inflammation. On the other hand, IRF4, along with nearly all other transcription factors, shows a positive correlation with pro-inflammatory genes. LINC01132, a down-regulated long non-coding RNA implicated in NF-kB signaling, exhibits a negative correlation with ATF3, SPIB and IKZF1. Interestingly, both of genes and TF presents sometimes at least one differentially methylated CpG in their promoter region. Along with miRNA and lncRNA, those epigenetic marks may explain the involvement of same TF in opposite pathways (pro and anti-inflammatory processes). This dualistic behavior could also be explained in variations among the TF combinations. To illustrate, IRF4, SPIB, and ATF3, which are among the most interconnected transcription factors within the entire network, demonstrate distinctive co-regulated gene clusters either independently or in concert (**supplementary figure 11**).

**Figure 5.**
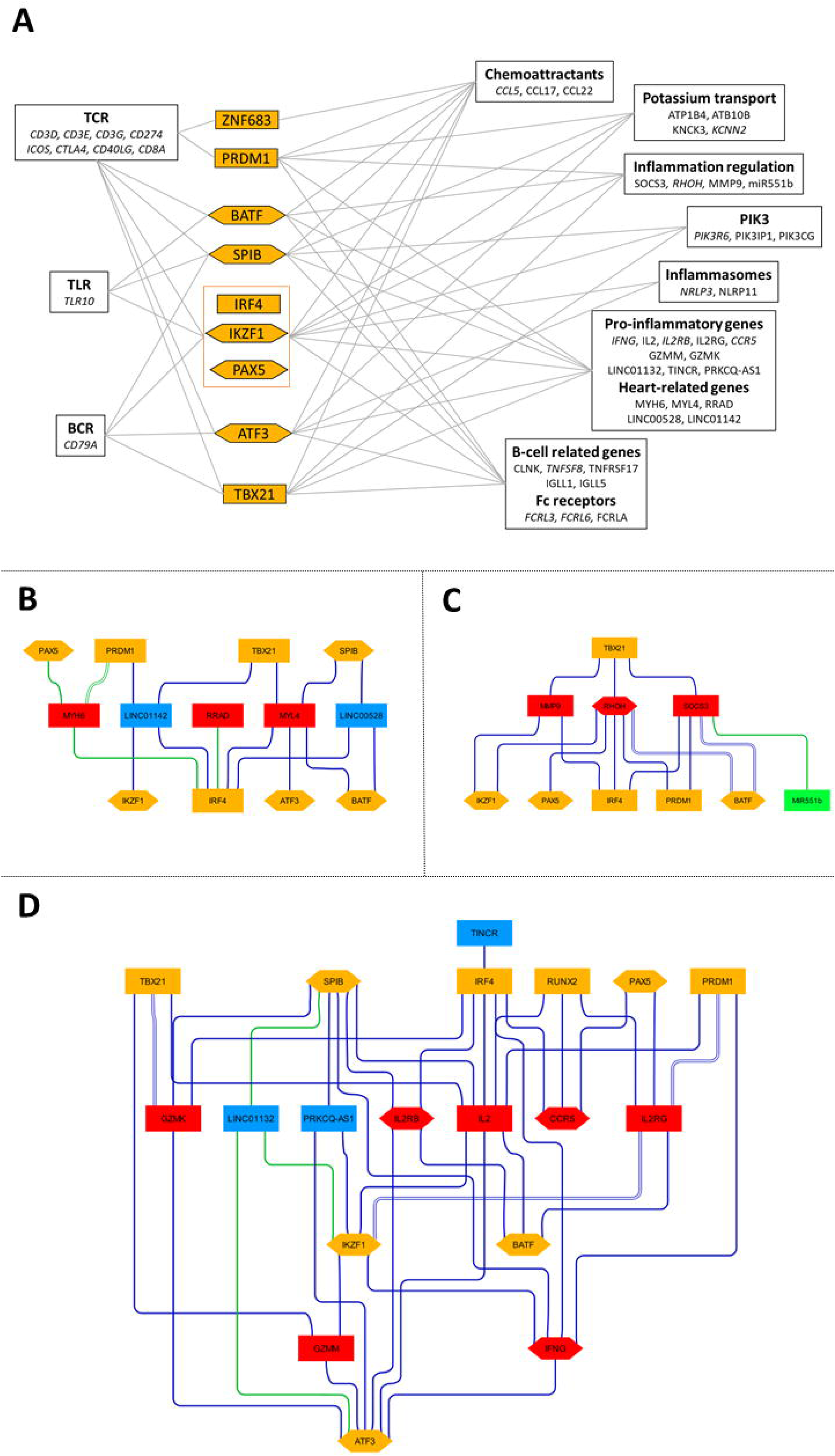
**(A)** Diagram illustrating the impact of top transcription factors on the biological processes influenced by genes within cluster 2. **(B)** Correlation network of the transcription factor targets genes involved in heart-related processes. **(C)** Correlation network of the transcription factor targets genes involved in inflammation regulation. **(D)** Correlation network of the transcription factor targets genes involved in inflammation. Nodes color: red: protein-coding genes, blue: lncRNA, green: miRNA, orange: transcription factor. Nodes shape: square: feature expression, hexagon: feature expression carrying at least one CpG selected with MOFA+ in its promoter. Edge line color: blue: positively correlated, green: negatively correlated. Edges line type: line: correlated in CCC network, dot: correlated in CTRL network, double line: correlated in both networks.

### TLR may regulate innate immune response

Considering the high convergence observed in BCR, suggesting a high-pressure selection on clonotype and therefore a continuous stimulation, we performed the same analysis on cluster 1 and 3, significantly enriched in BCR and TLR signaling. SPIC is the most highly connected transcription factor and appears to independently target numerous genes. To a lesser extent, it may also cooperate with FOSL1, a transcription factor involved in the regulation of inflammation (**supplementary figure 10B**). Focusing on potential origin of inflammation response, we have identified several genes involved in both innate (TLR, complement) and adaptive (BCR, TCR) immune system. 7 transcription factors showed correlations with at least one of these processes, with 6 of them associated with BCR genes. All those TF are correlated with inflammatory response. Interestingly, RUNX3, IRF8, SPIC and MYB are also correlated with immune regulation related genes (**figure 6A**). Looking at the resulting networks, SPIC is negatively correlated with TLR6, TLR8 as well as inflammasome (PYCARD) and mitochondria (CYP4F22) genes, but positively correlated with TLR7 (**figure 6B-6D**). While SPIC is negatively correlated with GAPT, LAIR1 and TNFAIP8L2, all associated to adaptative immune response regulation, it demonstrates positive correlations with PLD4 and GAPT, two genes linked to the regulation of innate immune responses (**figure 6E**). Similarly, it also exhibits a negative correlation with numerous genes linked to inflammation, including LINC01010, a target of MyD88 (**figure 6F**). In a lesser extent, ASCL2 and MYB, correlated with C2 gene, are also negatively correlated with some inflammatory genes. Interestingly, ASCL2 is negatively correlated with CCDC88B, required for T-cell pathway, in control network. Considering the DNA methylation observed in both ASCL2 and CCDC88B promoter, we can hypothesize that methylation may influence the regulation of CCDC88B by ASCL2. All these results tend to demonstrate the involvement of SPIC in innate immune regulation, probably through TLR7 stimulation.

**Figure 6.**
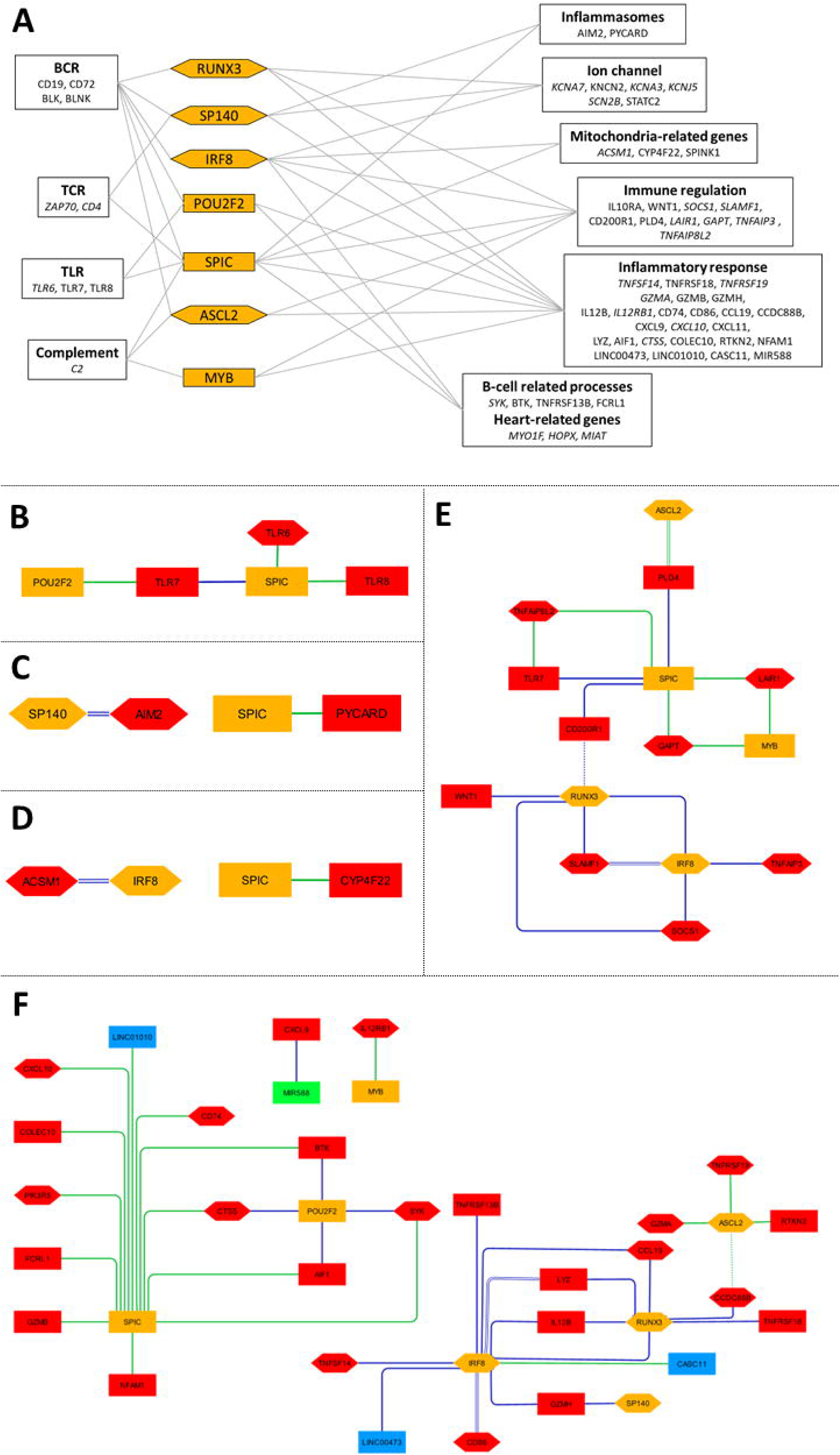
**(A)** Diagram illustrating the impact of top transcription factors on the biological processes influenced by genes within cluster 2. **(B)** Correlation network of the transcription factor targets genes involved in TLR signaling. **(C)** Correlation network of the transcription factor targets genes involved in inflammasomes. **(D)** Correlation network of the transcription factor targets genes associated to mitochondria process. **(E)** Correlation network of the transcription factor targets genes involved in immune response regulation. **(F)** Correlation network of the transcription factor targets genes involved in inflammatory response. Nodes color: red: protein-coding genes, blue: lncRNA, green: miRNA, orange: transcription factor. Nodes shape: square: feature expression, hexagon: feature expression carrying at least one CpG selected with MOFA+ in its promoter. Edge line color: blue: positively correlated, green: negatively correlated. Edges line type: line: correlated in CCC network, dot: correlated in CTRL network, double line: correlated in both networks.

### Epigenetic regulation at the heart of CCC pathogenic process

Considering the high number of differentially methylated genes and transcription factors observed in cluster 1, 2 and 3, a stringent filter was applied to the network, selecting only protein coding and transcription factors correlated at both expression and methylation level in one phenotype, or inversely correlated in both phenotypes. By this way, we ensure that the methylation may be responsible of the differential expression observed. As lncRNA and miRNA may not be affected by methylation in gene promoter, we conserved all these correlations. The resulting network highlight the potential impact of RUNX3 on pro-inflammatory genes expression, as well as Th2 T-cell repression through CCD8 (**figure 7**). Moreover, some miRNA may have a crucial role in the exacerbate inflammation occurring in CCC, such as MIR588, positively correlated with BCL2L15, RAB3C and CXCL9. On the opposite, the lack of miRNA binding may enhance inflammatory process, like MIR375, known to reduce apoptosis in cardiac cell, and correlated with IRF4 in control network only. Given the high number of lncRNA correlated with IRF4, it is plausible that its activity is sequestered or modulated by one of them. It is notable that IFNG also carried at least one differentially methylated CpG in its promoter and may be up regulated by PRDM1 transcription factor. While this analysis did not reveal many TF-TF potential interactions (based on both expression and methylation correlations), many TF selected by MOFA+ have at least one CpG differentially methylated in their promoter. The methylation level may not gradually affect the binding but act as a switch. Looking on all the TF-TF correlations, we observed that PAX5, IRF4 and ATF3 are the top-connected TF, but IRF4 is the only one that do not carry any differentially methylated CpG site (**supplementary figure 12**). Although alterations in methylation may not definitively signify whether the correlated transcription factor will or will not bind to this promoter in CCC, it remains interesting to explore this possibility for a deeper understanding of the pathogenic process.

**Figure 7.**
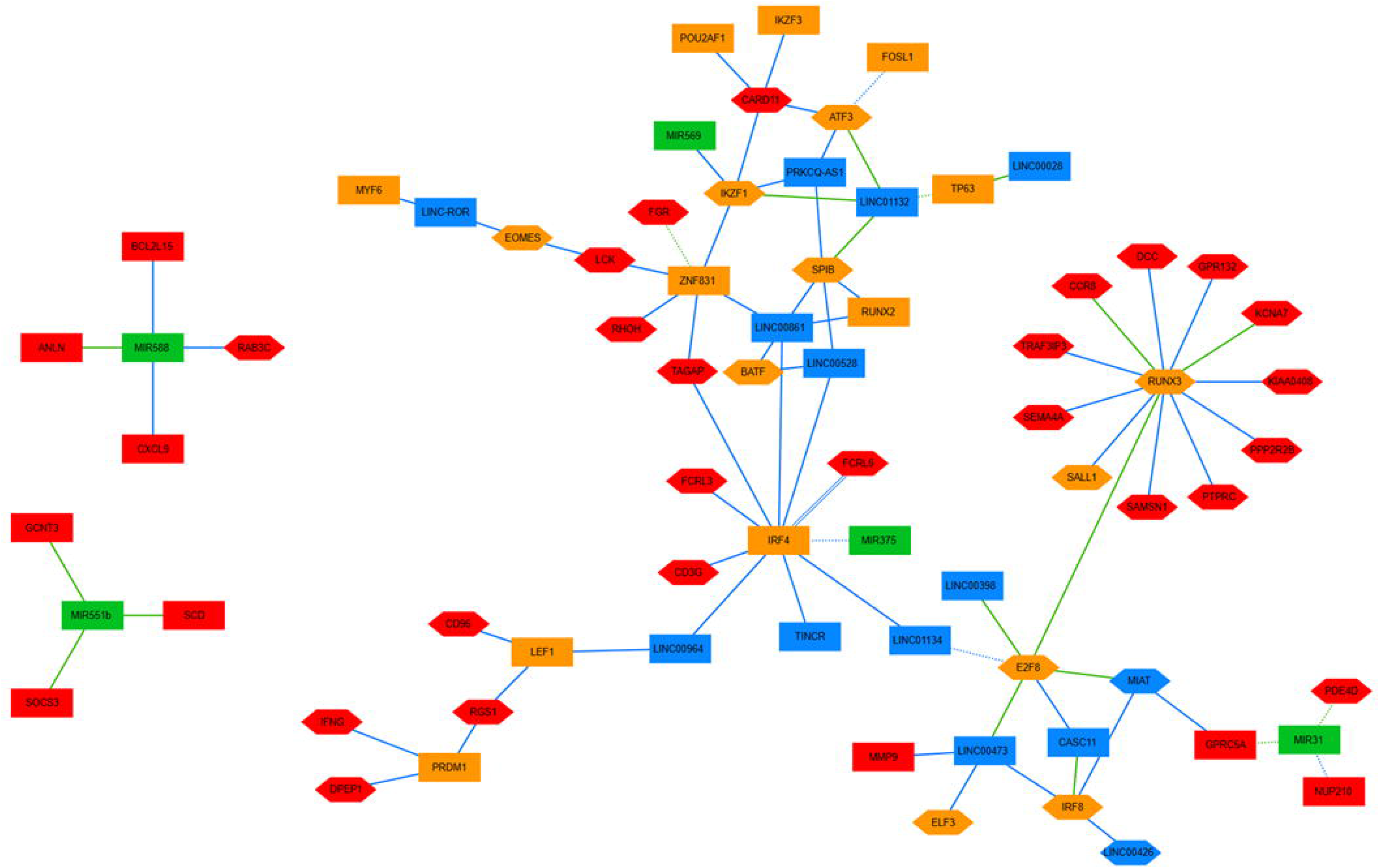
Correlation network of genes potentially affected by at least one CpG selected with MOFA+ in its promoter of one ncRNA. Nodes color: red: protein-coding genes, blue: lncRNA, green: miRNA, orange: transcription factor. Nodes shape: square: feature expression, hexagon: feature expression carrying at least one CpG selected with MOFA+ in its promoter. Edge line color: blue: positively correlated, green: negatively correlated. Edges line type: line: correlated in CCC network, dot: correlated in CTRL network, double line: correlated in both networks.

### MOFA factor discriminate 5 of the 7 CCC patients

Beyond the shared variability observed across all CCC patients compared to healthy controls, a secondary multi-omics integration was conducted with the purpose of examining individual patient differences, with the aim of determining whether the disease may have diverse origins. 4 factors carrying at least 1% of the variability in the data were used to discriminate 5 of the 7 CCC patients (**figure 8A**). Features with positive or negative weights in these factors are enriched in Reactome pathways related to mitochondrial function (respiratory electron transport, complex I biogenesis), innate immune response (complement cascade, Toll Like Receptor Cascade, interferon signaling), nervous system or muscle contraction (**supplementary figure 13).** As all these processes are directly linked to chronic chagasic cardiomyopathies, the discriminative features obtain through the different factors may be involved in the pathogenesis. Furthermore, except for factor 3, the top-features of the factors are only partially known in ChagasDB (**figure 8B**).

**Figure 8.**
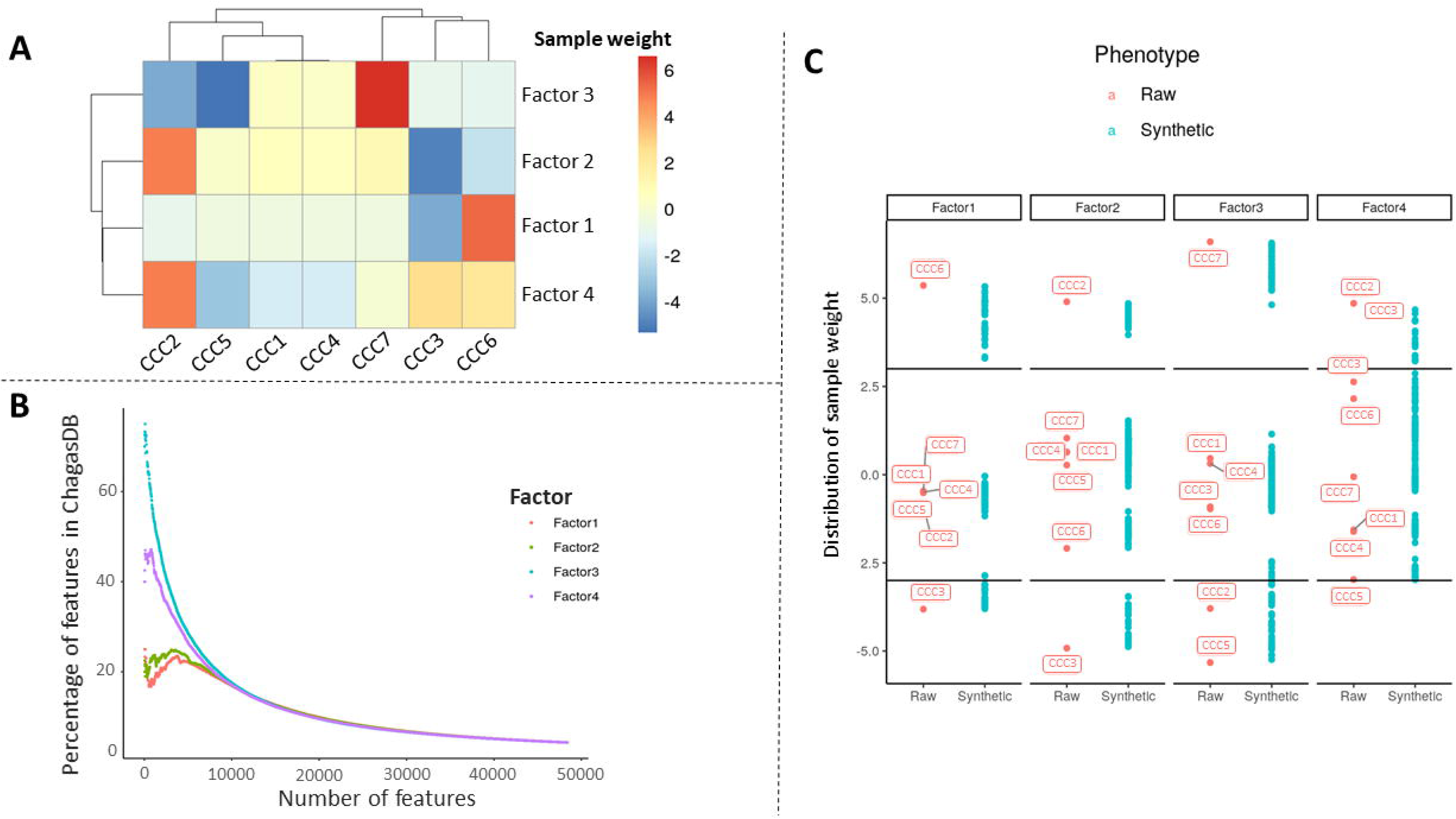
**(A)** Clustering of CCC samples according to their contribution in each factor. **(B)** Distribution of CCC samples contribution in each factor. **(C)** Percentage of features present in each factor within ChagasDB, arranged based on their respective weights in each factor.

### CCC patients present specific expression profiles

In order to understand the specificity of each patient, the 50% of features that contributed most to the various factors were selected (**supplementary table 7)**. They were then associated with each sample according to their contribution to each factor. Considering the synthetic samples associated to each raw samples, 5 clusters of samples were selected, assigned to CCC6 (n=20), CCC3 (n=19), CCC2 (n=19), CCC7 (n=20) and CCC5 (n=17) (**figure 8C**). The 5 sample clusters bring together genomic, trancriptomic and methylomic data. However, with the exception of methylation sites, few of these are sample-specific (**table 2**). This is particularly the case for the CCC2 and CCC7 clusters, where a small number of protein-coding genes have been selected. Three samples stand out from the rest with specific features involved in precise biological processes: CCC6 (mitochondrial alterations, notably complex I), CCC3 (activation of T-CD4 lymphocytes and the Wnt pathway) and CCC5 (Toll-like receptor signaling activity) (**figure 9**). As CCC3 and CCC6 are the only samples not imputed characterized by specific biological processes, further analysis was conducted only on these samples.

**Figure 9.**
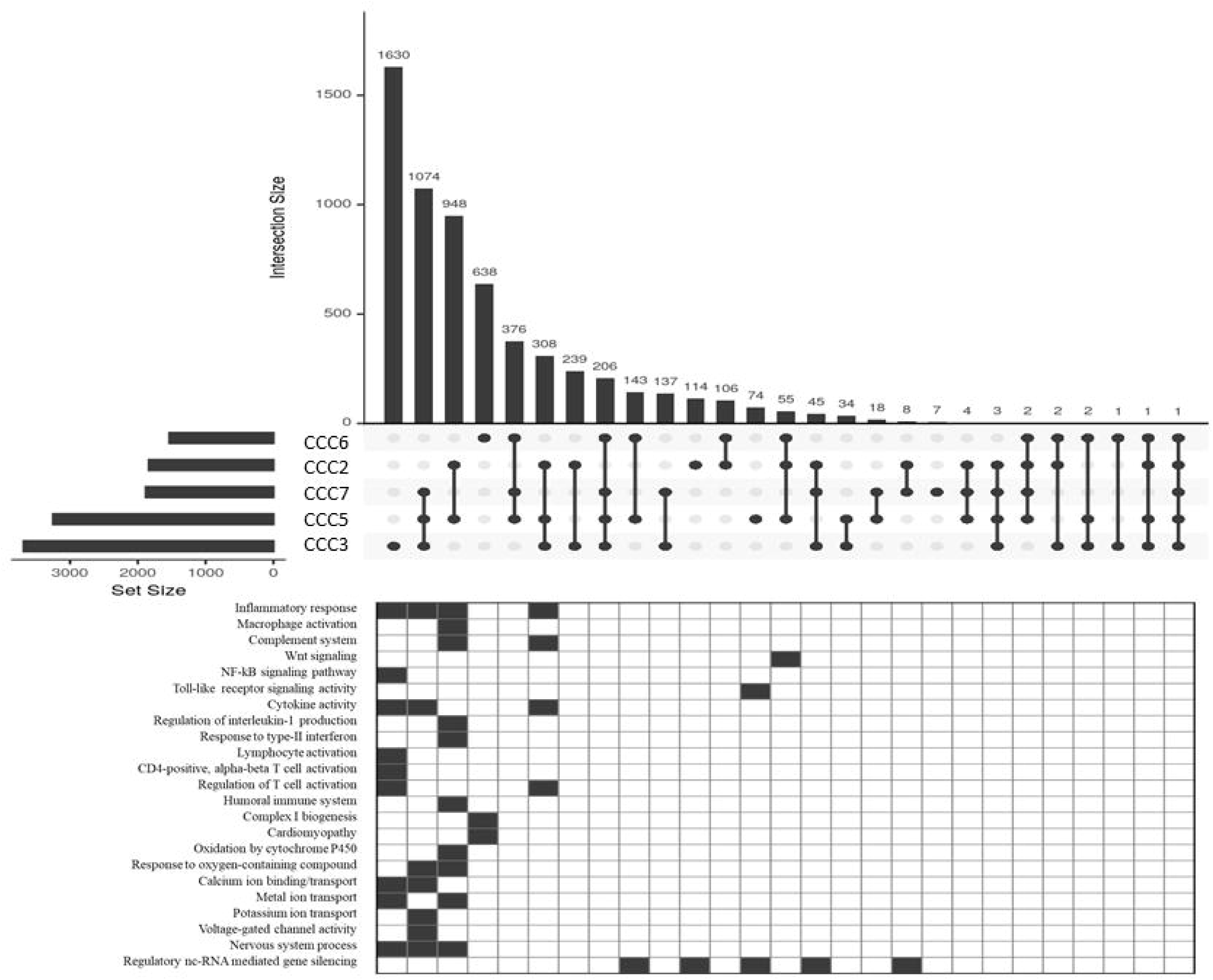
Upset plot of the common expressed features between the 5 discriminates CCC samples, and corresponding enriched biological process. The horizontal bars represent the total number of features to each sample cluster, while the vertical bars represent the number of features unique to the different comparisons.

### Mitochondria mutation as a cofactor of CCC pathogenesis

Interestingly, 9 mitochondrial mutations were specifically assigned to CCC6 cluster. Among them, MT-ND1 3604 CTAG > C was the top-weighted mutation in factor 1 (weight = 3.43) compared to others (weight < 1.55). Regarding the differential gene expression between the samples carrying this mutation and the other, 2 miRNAs present significant differential expression (abs(log2FC) >= 1.5 & adjusted p-value <= 0.05): miR-369-5p and miR-591. Therefore, this mutation may be involved in the difference of expression observed with these 2 miRNAs.

### IRF4 exacerbation as a marker of CCC inflammation

As before, a correlation network was build using the features only associated to CCC3 sample. The resulting network illustrates higher expression on IRF4 transcription factor, as well as some ncRNA including MIR31, TINCR, IRF4, MIR146a, MIR141 and MIR1224. While half of the observed correlations are shared by all samples, CCC3 exhibits a unique miRNA profile, potentially up-regulated genes involved in CCC pathogenesis such as NLRP3 or IL21R (**figure 10**). Interestingly, two miRNAs are correlated to IRF4 and may enhance its expression, resulting in a worse B/T cell inflammatory response.

**Figure 10.**
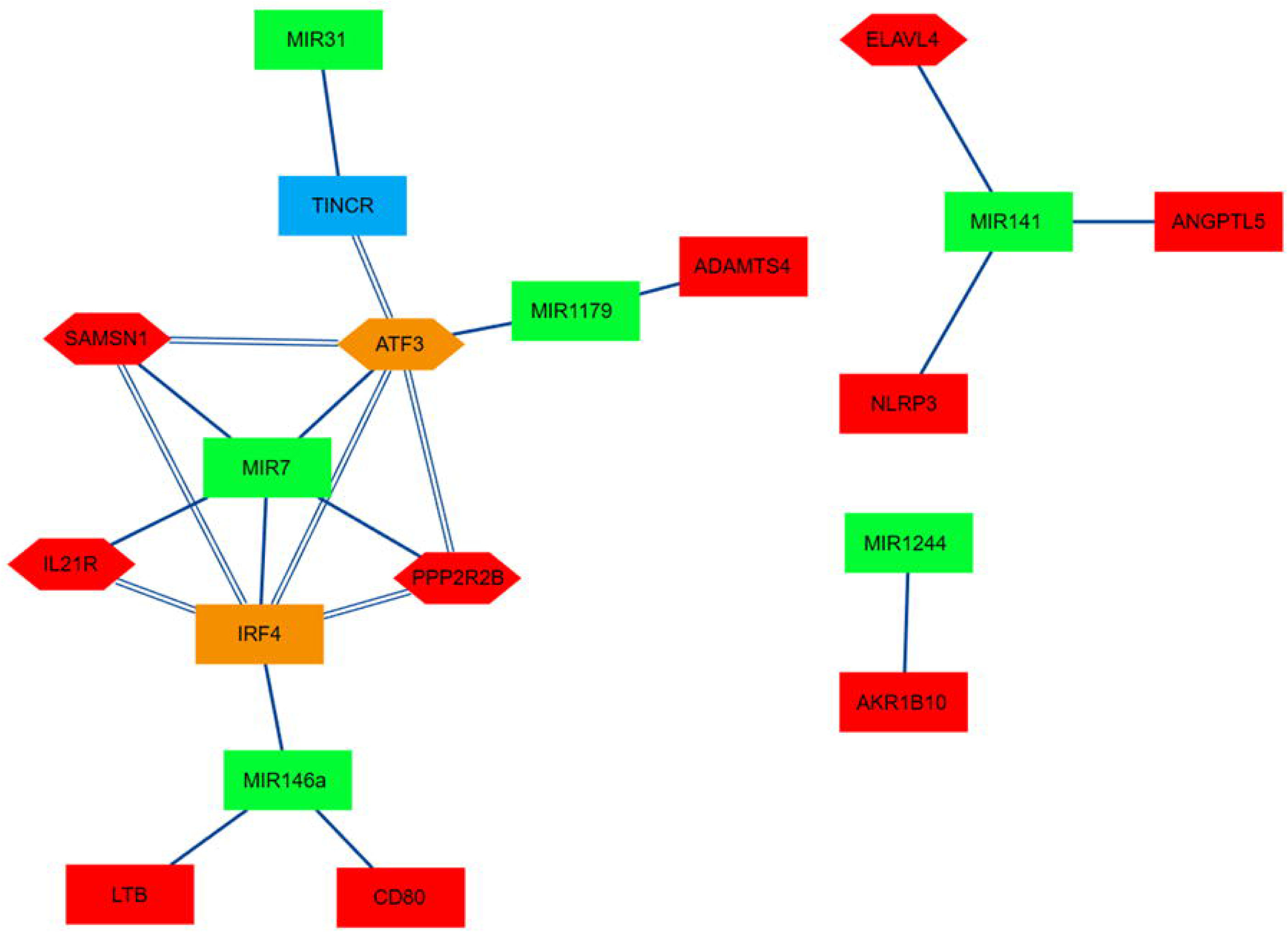
Correlation network of genes only associated to CCC3 cluster. Nodes color: red: protein-coding genes, blue: lncRNA, green: miRNA, orange: transcription factor. Nodes shape: square: feature expression, hexagon: feature expression carrying at least one CpG selected with MOFA+ in its promoter. Edge line color: blue: positively correlated, green: negatively correlated. Edges line type: line: correlated in CCC network, dot: correlated in CTRL network, double line: correlated in both networks.

### Both innate and adaptative immune response leads to CCC development

To confirm our previous findings, we conducted a comparative analysis of key features across all CCC samples, with a particular focus on CCC3 and CCC6 (**figure 11**). Notably, CCC3 and CCC6 exhibited elevated levels of both SPIC and TLR7. However, CCC6 displayed a higher expression of TLR8, BLNK, and CD19. In contrast, CCC6 exhibited higher expression levels of TLR10, IFNG, IRF4, NRLP3, and CD8A. Examining the BCR/TCR repertoire, CCC6 stood out as having one of the highest numbers of clonotypes. While this may suggest a greater presence of B/T cells in the CCC myocardium, these results may also indicate a more robust humoral response in CCC6, leading to an elevated expression of IFNG. Furthermore, CCC6 exhibited increased expression of both TLR7 and TLR8, which are endosomal TLRs, along with genes related to B cells, underscoring an internal response. While BCR convergence was observed in all samples, both CCC3 and CCC6 resulted in heart tissue damage through distinct processes.

**Figure 11.**
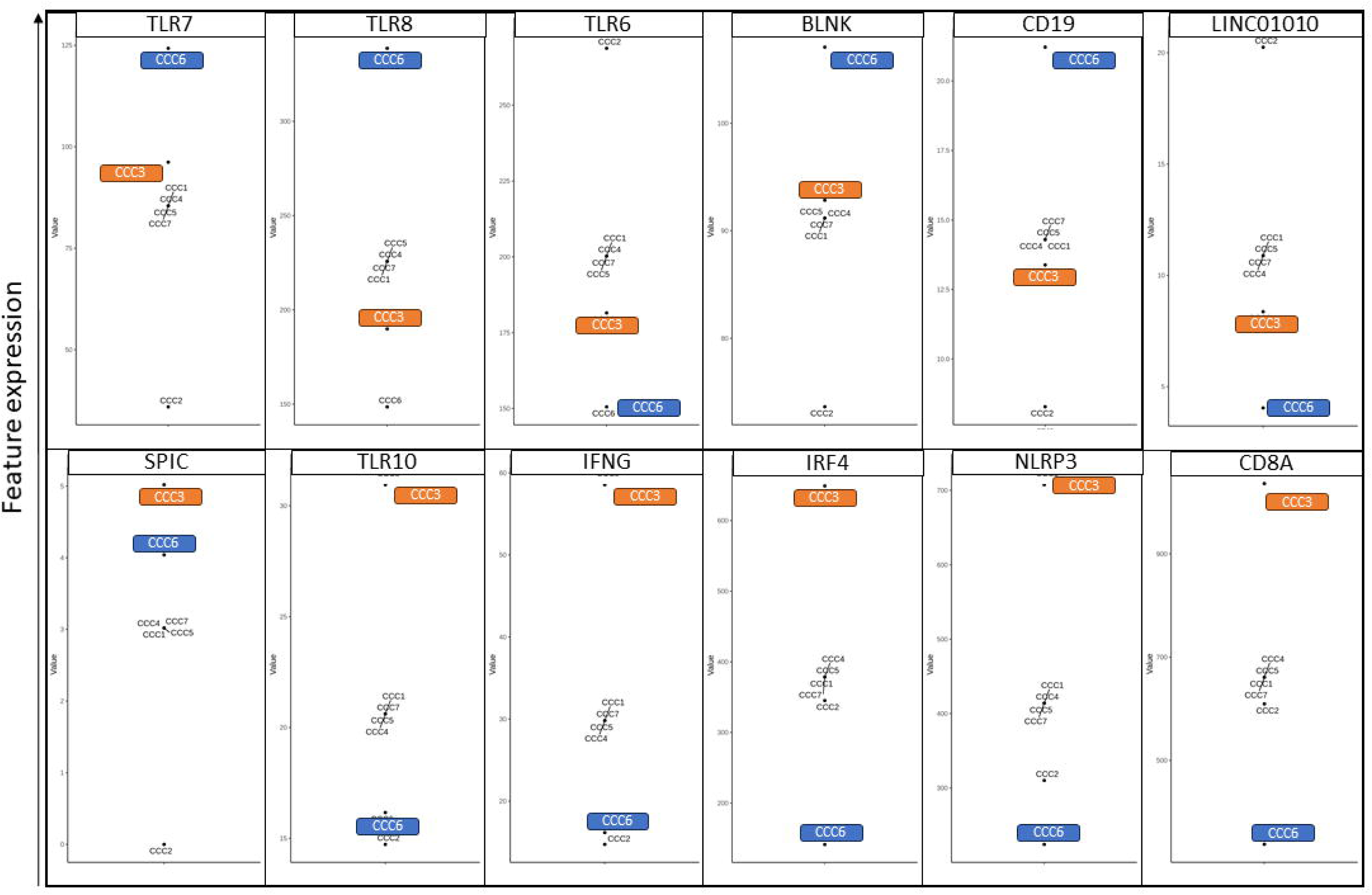
Expression level of the identified features of interest in the previous results, with CCC3 in orange and CCC6 in blue.

## Discussion

In this study, we explored potential factors influencing gene expression in CCC at different omics levels. This multi-omics integration was conducted in a small and incomplete dataset, primarily because of challenges in obtaining human heart samples. To counteract these difficulties, the decision was made to impute the missing values using the entire multi-omics dataset. This approach produced convincing results, with the elimination of samples having minimal impact on the characteristics selected following MOFA analysis. The choice to avoid existing methods for creating synthetic samples, such as SMOTE and KNNOR, was influenced by the complexity of processing different omics data. These methods rely on k-nearest neighbors to generate synthetic samples, which can be particularly challenging in a multi-omic dataset where each omics has different number of features. Our approach has overcome these difficulties by avoiding the need for such neighbor selection. In single-omic datasets, the results are very close to those obtained with SMOTE, which reinforces the preference for this solution.

Based on genomic, transcriptomic and methylomic dataset, the multi-omics integration has highlighted 1,214 genes, 2,066 methylation site and 58 miRNA discriminating healthy controls to CCC patients. Moreover, 488/1272 expressed features (46%) were never associated to this disease, according to ChagasDB (9). The genes were mostly associated to immune-related processes, mostly involved in inflammatory response and Th1 cell activation. The involvement of Th1-cell related genes in CCC has already been largely demonstrated in previous study (10,11,14).

In order to understand the relations between these features, two correlation networks were built, each specific to one phenotype. The same features exhibit contrasting behaviors between healthy control and CCC patient networks. In CCC, three regulatory clusters were observed, including genes involved in T-cell/TCR processes, or BCR and TLR pathways. We have identified several transcription factors correlated with genes involved in opposite biological processes, notably pro- and anti-inflammatory response. Among them, IRF4 may act with different TF, resulting in different functions. IRF4 is a transcription factor mostly produced by B cell after BCR stimulation by antigens and co-stimulatory signal with T-cell (37) but also in T cell after TCR stimulation. In our data, IRF4 is correlated with both TCR and BCR genes. In vitro, IRF4 overexpression leads to inflammasome inhibition (38), while NLRP3 is overexpressed in CCC samples, suggesting a default in inflammatory regulation. Interestingly, its impact on the immune response depends on its interactors and the cell type in which it is produced (39). Notably, in T-cell, it interacts with BATF, and in B cell, with SPIB. In Treg specially, it has been shown to interact with FOXP3. In our data, IRF4 is correlated with FOXP3 in CTRL network only, and with BATF (AP-1 family) in CCC network. Recently, Arnold PR. et al demonstrate that BATF3, a TF of AP-1 family, partnering with IRF4 to repress FOXP3, and so Treg production (40). On the opposite, both IRF4 and BRDM1 are correlated with CTLA4, a Treg marker, and with SOCS3, a regulator of STAT3 and downregulated in CCC. The reduction of SOCS3 gene expression was associated to IL-10 expression and Th2 profile induction (41). SOCS3 is also negative correlated with miR-551b, an activator of STAT3 (42). In our data, STAT3 is under-expressed in CCC compared to CTRL, possibly resulting from a lack of activation by miR-551b. Furthermore, various studies have linked IRF4 to numerous transcription factors observed in CCC, such as RUNX3 (43), PAX5 (44), IKZF1 (45), TBX21, and EOMES (46). Our analysis suggests that RUNX3 plays a pivotal role in activating the inflammatory response, potentially influenced by DNA methylation within its target promoter region. A prior study from our research predicted that both PAX5 and IRF4 may bind to the RUNX3 promoter, with their binding potentially regulated by differences in methylation levels (11). Additionally, our in vitro experiments demonstrated that methylation of the RUNX3 promoter impacts its expression (14). Within the CCC network, we observed that CCC6 sample is characterized by elevated IRF4 expression, along with increased levels of IFN-γ and the NLRP3 inflammasome. Considering the CD8A and CD4 gene markers, as well as a high number of TCR clonotypes, it is plausible that IRF4 is associated with an intensified T-cell response, leading to cardiac damage. In this line, IRF4 exhibits correlations with LINC00964 and LINC00528, both of which have already been implicated in heart diseases (47,48). MIAT, a lncRNA previously linked to CCC (13) and present in our network, is also overexpressed in CCC6 patient. Collectively, these findings strongly indicate that IRF4 serves as a key regulator of the inflammatory response.

Our analysis has also revealed some TLR overexpressed in CCC patients, including TLR6, TLR7, TLR8 and TLR10. TLR10 is an endosomal TLR mainly located in B cell (49), potentially activated by double-stranded DNA. Although little known, TLR10 seems to differ from other TLRs in its anti-inflammatory action (50). However, in our data, TLR10 is correlated with pro-inflammatory genes, TLR4 and BCR-related genes (CD79A, BANK1), suggesting a dual function of this TLR. On the other hand, TLR7 is correlated with SPIC, and both are linked to immune regulation genes. In vitro and in vivo experiments highlights the SPIC activation through the TLR/NF-kB pathway (51), inducing the production of anti-inflammatory cytokines. In this line, SPIC is also correlated to IL-4 and IL-4I1, a cytokine involved in Th2 differentiation (52), as well as FRZB, a regulator of Wnt pathway (53,54). We can hypothesize here that SPIC may play an inflammatory regulation process, while not sufficient to reduce the inflammation in CCC. It is interesting to note that both TLR7 and SPIC are up regulated in CCC3 and CCC6 patients. On the other hand, TLR8 is positively correlated with inflammatory genes, and up regulated in CCC6 sample. TLR8 is an endosomal TLR activated by single-stranded DNA. Considering that *T.cruzi* DNA is single-stranded, it may be directly activated by resting amastigotes in cardiomyocytes. However, CCC6 samples present mitochondrial dysfunctions, potentially due to several mitochondria mutations. Mitochondrial dysfunctions are known to participate to CCC pathogenic processes. The accumulation of defective mitochondria may lead to mitophagy (55), resulting in the generation of mitochondria DAMP, which can activate endosomal TLR (56). While mtDNA is double-stranded, RNA:DNA hybrids form produced during the transcription could be recognized by TLR (57).

Regarding specifically at CCC6 mtDNA genotype, the rs28647976 mutation, a missense variant in MT-ND1, a gene involved in complex I, was associated to miR-369-5p and miR-591. The overexpression of miR-369 conduct to a reduction of inflammation in rat (58). Moreover, miR-369-5p target DNMT3 gene, conducting in reduction of cardiac fibroblasts proliferation (59). The miR-369-3p is associated to a reduction of NO production (60) and inflammasome pathway (61). In CCC context, may be attempting to mitigate the oxidative stress that is exacerbated by both CCC pathogenic process and MT-ND1 mutation, and therefore be an interesting therapeutic target. The identification of specific mutations on some samples are in line with the family-specific mutations previously identified (62).

While different pathogenic pathways have been identified in different CCC patients, the BCR repertoire analysis support the existence of specific selection of BCR light chains in the absence of the pathogen in all CCC samples. The convergence towards sequences common to all the patients studied indicates the possibility of the establishment of B cell autoreactivity against similar host proteins, in line with previous observations in humans and mice (8). Interestingly, IRF4 is required to perform the light chain rearrangements (63). In addition, some study reveals the involvement of both BCR and TLR in the development of auto-immune disease (64,65). Our multi-omics integration identified several features known to be involved in autoimmune disease, including FCRL5, one of the top discriminative features (66). The hypothesis of an autoimmune reaction occuring in CCC heart tissue has already been supported by different studies. For example, T-cell obtains from CCC patients reacts after stimulation by T.cruzi derived peptides, like the B13 protein (67). Considering the absence of TCR public repertoire in our sample, the specificity of TCR, and the resulting T-cell response, may be variable on the different samples. This result could also be due to an insufficiant sequencing depth, and BCR/TCR specific analysis should be performed.

To conclude, this analysis has identified numerous regulatory elements probably involved in CCC pathogenesis. While all CCC samples present a similar B-cell response, some of them develop specific process, leading to the heart cardiomyopathy. Considering the opposite roles mediated by TF in different cell types, further analysis at single-cell levels are required to conclude about the importance of anti-inflammatory processes observed in CCC, and their lack of efficiency. Nonetheless, the observations conducted in this study could help, in the future, to develop effective treatments depending on the divergent reactions of each patient.

## Data Availability

All data produced in the present study are available upon reasonable request to the authors

## Acknowledgments

Center de Calcul Intensif d’Aix-Marseille is acknowledged for granting access to its high-performance computing resources.

## Tables

**Table 1.** Human heart tissue available through the different omics.

**Table 2.** Top 50% features selected in each factor and associated to each sample. The total number of features, and the number of not shared features across the samples are displayed.

## Supplementary materials

**Supplementary Figure 1.** (A) Statistical power estimation with the increasing number of samples per omic. (B) Alluvial plot of the common selected features with the increasing number of samples.

**Supplementary Figure 2.** Visualization of the dimension 5 and 6 of PCA analysis performed, for each phenotype (BRCA and LUAD) on raw and synthetic samples generated with bootstrapping, SMOTE, KNNOR or barycenter methods (red: raw samples, blue: synthetic samples).

**Supplementary Figure 3.** Comparison of the selected features after different multi-omics integration tools. (A) Venn diagram between MOFA+, MCIA and JIVE. (B) Upset plot comparing the features in common between MOFA+, MCIA, JIVE and MixOmics integration. The horizontal bars represent the total number of features per method, while the vertical bars represent the number of features unique to the different comparisons.

**Supplementary Figure 4.** Comparison of the selected features after different iteration, removing each type one raw sample in one omic. (A) Jaccard index, (B) minimal overlap.

**Supplementary Figure 5.** Evaluation of the 7-factor selected with CTRL and CCC samples. (A) Percentage of variance explained per omic and per factor. (B) Correlation between each factor.

**Supplementary Figure 6.** Top enriched Reactome pathway in factor 1. Red: positive features; blue: negative features.

**Supplementary Figure 7.** Comparison between CTRL and CCC network. (A) Distribution of the correlation between paired features in one phenotype, when the correlations are high (|r| >= 0.8) in the other phenotype. (B) Venn diagram of the conserved features in each network. (C) Venn diagram of the pairwise correlation between pairs of features in each network. Red: CCC; blue: CTRL.

**Supplementary Figure 8.** Clonotypes analysis of T cell receptor. (A) Abundance of TCR clonotypes per phenotype. (B) Distribution of the number of samples sharing public clonotypes per phenotype: 1) All samples, 2) Control samples, 3) CCC samples.

**Supplementary Figure 9.** Frequency distribution of the V genes associated with the BCR sequences. (A) Cluster 1, (B) Cluster 2, (C) Cluster 3, (D) Cluster 4, (E) Cluster 5, (F) Cluster 6, (G) Cluster 7, (H) Cluster 8, (I) Cluster 9, (J) Cluster 10, (K) Cluster 11.

**Supplementary Figure 10.** Upset-plot of the top-10 connected TF in (A) cluster 2 and (B) clusters 1 and 3. The horizontal bars represent the total number correlated features to each TF, while the vertical bars represent the number correlated features unique to the different comparisons.

**Supplementary Figure 11.** Correlation network of IRF4 (red circle), SPIB (blue circle) and BATF (green circle) transcription factor targets genes. Nodes color: red: protein-coding genes, blue: lncRNA, green: miRNA, orange: transcription factor. Nodes shape: square: feature expression, hexagon: feature expression carrying at least one CpG selected with MOFA+ in its promoter. Edge line color: blue: positively correlated, green: negatively correlated. Edges line type: line: correlated in CCC network, dot: correlated in CTRL network, double line: correlated in both networks.

**Supplementary table 1.** List of BRCA and LUAD samples used from the TCGA database.

**Supplementary table 2.** Features weight in factor 1 obtained with MOFA+ CTRL/CCC integration.

**Supplementary table 3.** Type and number of features involved in each correlation network.

**Supplementary table 4.** Pairwise correlation used to build the CTRL and CCC network. The features having a methylation site in their promoter regions and the known interaction in public databases are noticed.

**Supplementary table 5.** Clustering assignment of protein coding genes and regulatory features.

**Supplementary table 6.** Type and number of features associated to each cluster.

**Supplementary table 7.** Features weight in factors 1-4 obtained with MOFA+ CCC integration.

**Supplementary table 8.** Pairwise correlation used to build the CCC3 network. The features having a methylation site in their promoter regions and the known interaction in public databases are noticed.

