## Supplementary material for "Exploring global and specific pathogenic mechanisms in Chronic Chagas Cardiomyopathy through multi-omics integration": Supplementary figure 4.pptx

### Slide 1
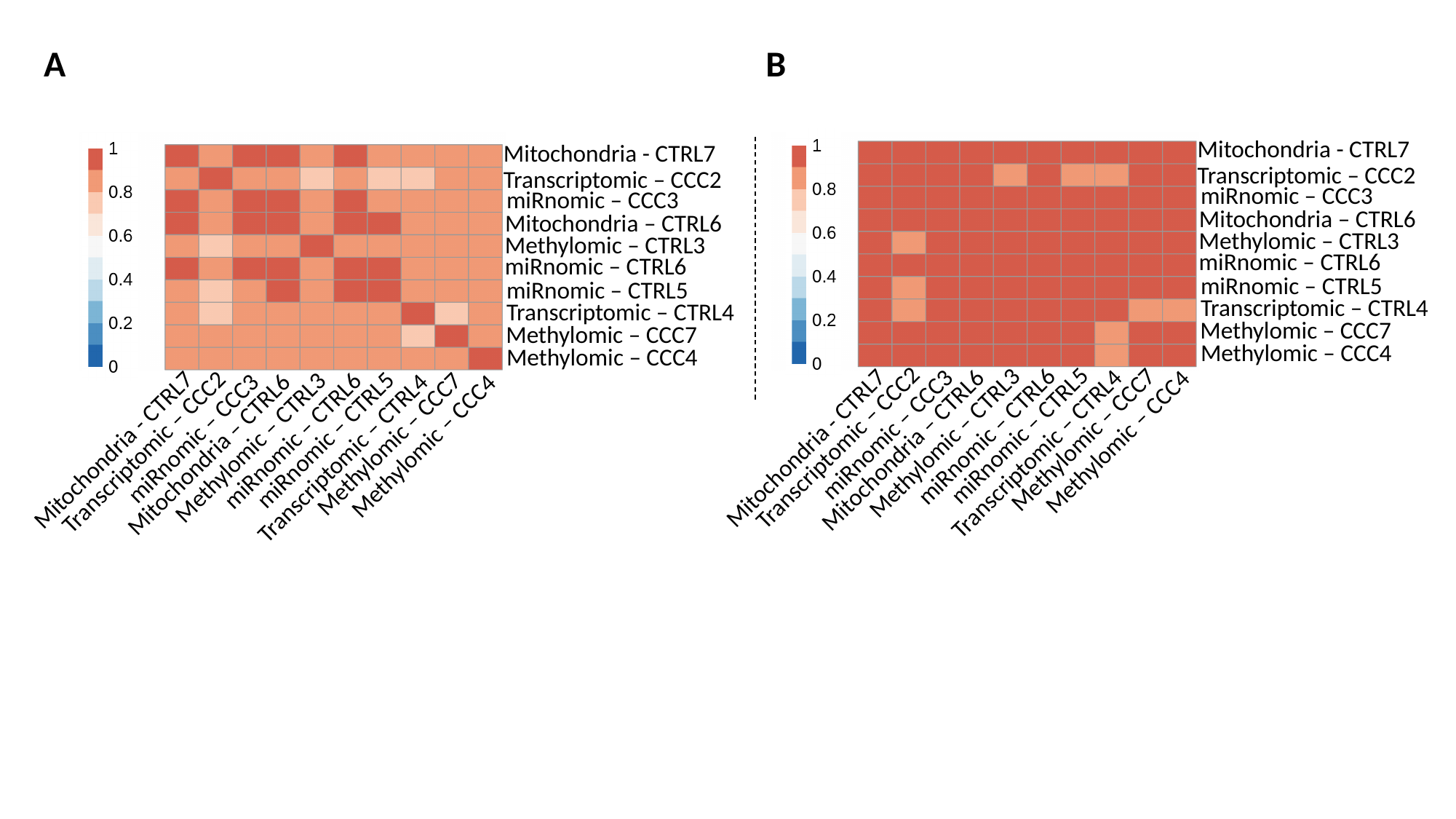

A
B
Mitochondria - CTRL7
Mitochondria - CTRL7
Transcriptomic – CCC2
Transcriptomic – CCC2
miRnomic – CCC3
miRnomic – CCC3
Mitochondria – CTRL6
Mitochondria – CTRL6
Methylomic – CTRL3
Methylomic – CTRL3
miRnomic – CTRL6
miRnomic – CTRL6
miRnomic – CTRL5
miRnomic – CTRL5
Transcriptomic – CTRL4
Transcriptomic – CTRL4
Methylomic – CCC7
Methylomic – CCC7
Methylomic – CCC4
Methylomic – CCC4
Transcriptomic – CCC2
miRnomic – CTRL5
Methylomic – CCC7
miRnomic – CTRL6
Methylomic – CTRL3
Mitochondria - CTRL7
Transcriptomic – CTRL4
miRnomic – CCC3
Mitochondria – CTRL6
Methylomic – CCC4
Transcriptomic – CCC2
Mitochondria - CTRL7
miRnomic – CTRL5
Methylomic – CCC7
miRnomic – CTRL6
Methylomic – CTRL3
Transcriptomic – CTRL4
miRnomic – CCC3
Mitochondria – CTRL6
Methylomic – CCC4
