## Supplementary material for "Exploring global and specific pathogenic mechanisms in Chronic Chagas Cardiomyopathy through multi-omics integration": Supplementary figure 5.pptx

### Slide 1
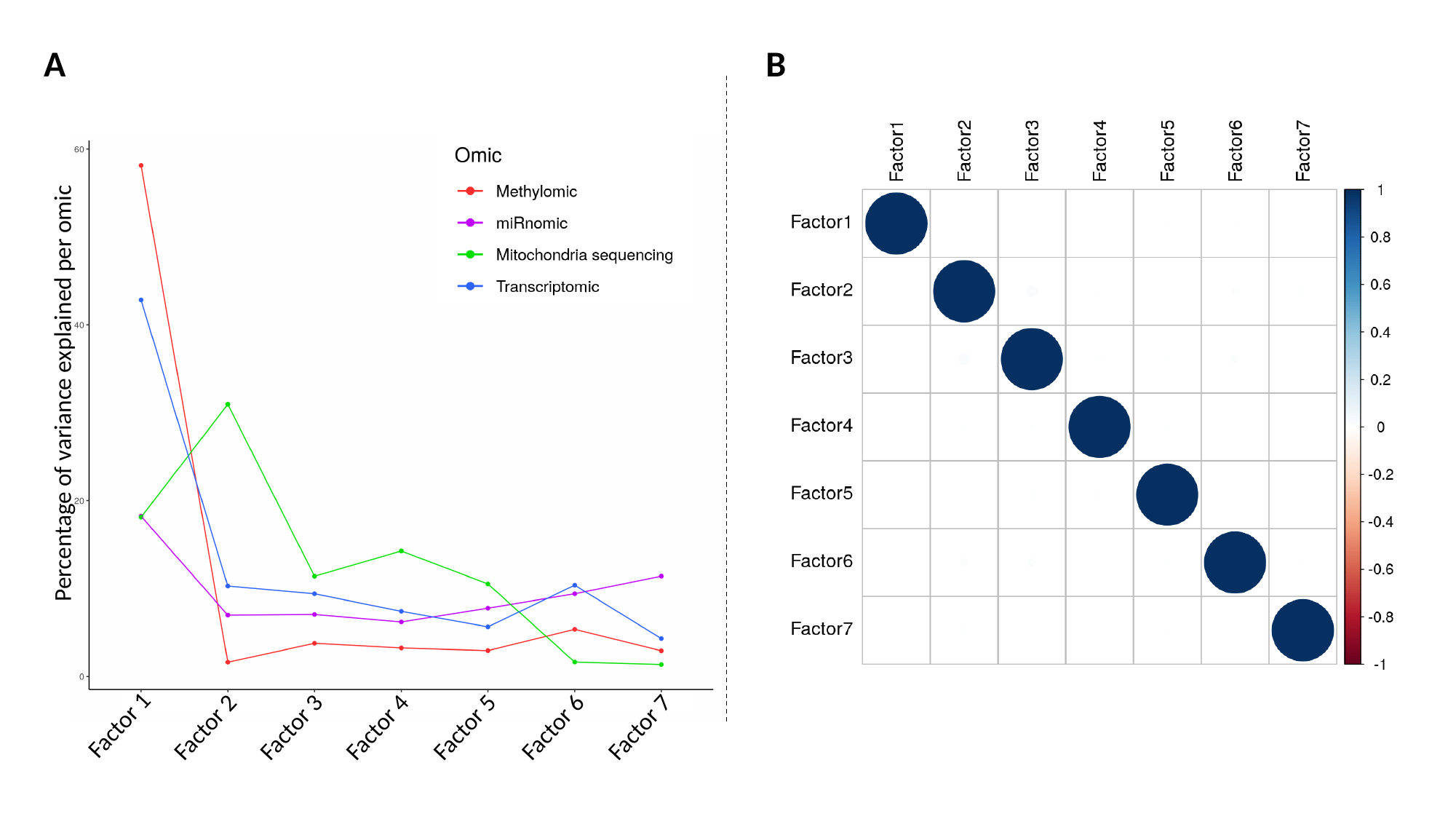

A
B
Percentage of variance explained per omic
Factor 1
Factor 3
Factor 4
Factor 5
Factor 6
Factor 2
Factor 7
