## Supplementary material for "Exploring global and specific pathogenic mechanisms in Chronic Chagas Cardiomyopathy through multi-omics integration": Supplementary figure 6.pptx

### Slide 1
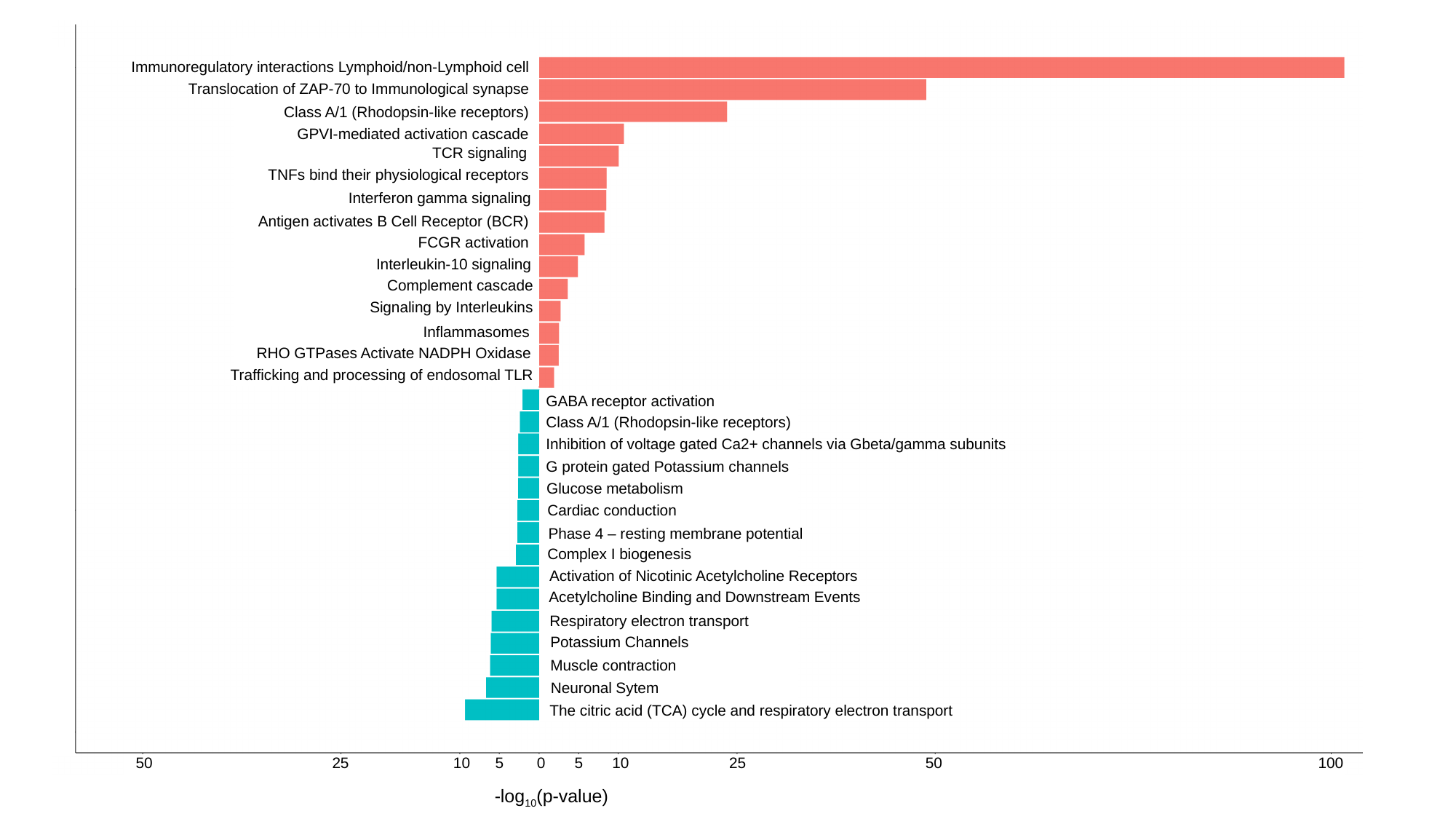

Immunoregulatory interactions Lymphoid/non-Lymphoid cell
Translocation of ZAP-70 to Immunological synapse
Class A/1 (Rhodopsin-like receptors)
GPVI-mediated activation cascade
TCR signaling
TNFs bind their physiological receptors
Interferon gamma signaling
Antigen activates B Cell Receptor (BCR)
FCGR activation
Interleukin-10 signaling
Complement cascade
Signaling by Interleukins
Inflammasomes
RHO GTPases Activate NADPH Oxidase
Trafficking and processing of endosomal TLR
GABA receptor activation
Class A/1 (Rhodopsin-like receptors)
Inhibition of voltage gated Ca2+ channels via Gbeta/gamma subunits
G protein gated Potassium channels
Glucose metabolism
Cardiac conduction
Phase 4 – resting membrane potential
Complex I biogenesis
Activation of Nicotinic Acetylcholine Receptors
Acetylcholine Binding and Downstream Events
Respiratory electron transport
Potassium Channels
Muscle contraction
Neuronal Sytem
The citric acid (TCA) cycle and respiratory electron transport
50 25 10 5 0 5 10 25 50 100
-log_10_(p-value)
