## Supplementary material for "Exploring global and specific pathogenic mechanisms in Chronic Chagas Cardiomyopathy through multi-omics integration": Supplementary figure 7.pptx

### Slide 1
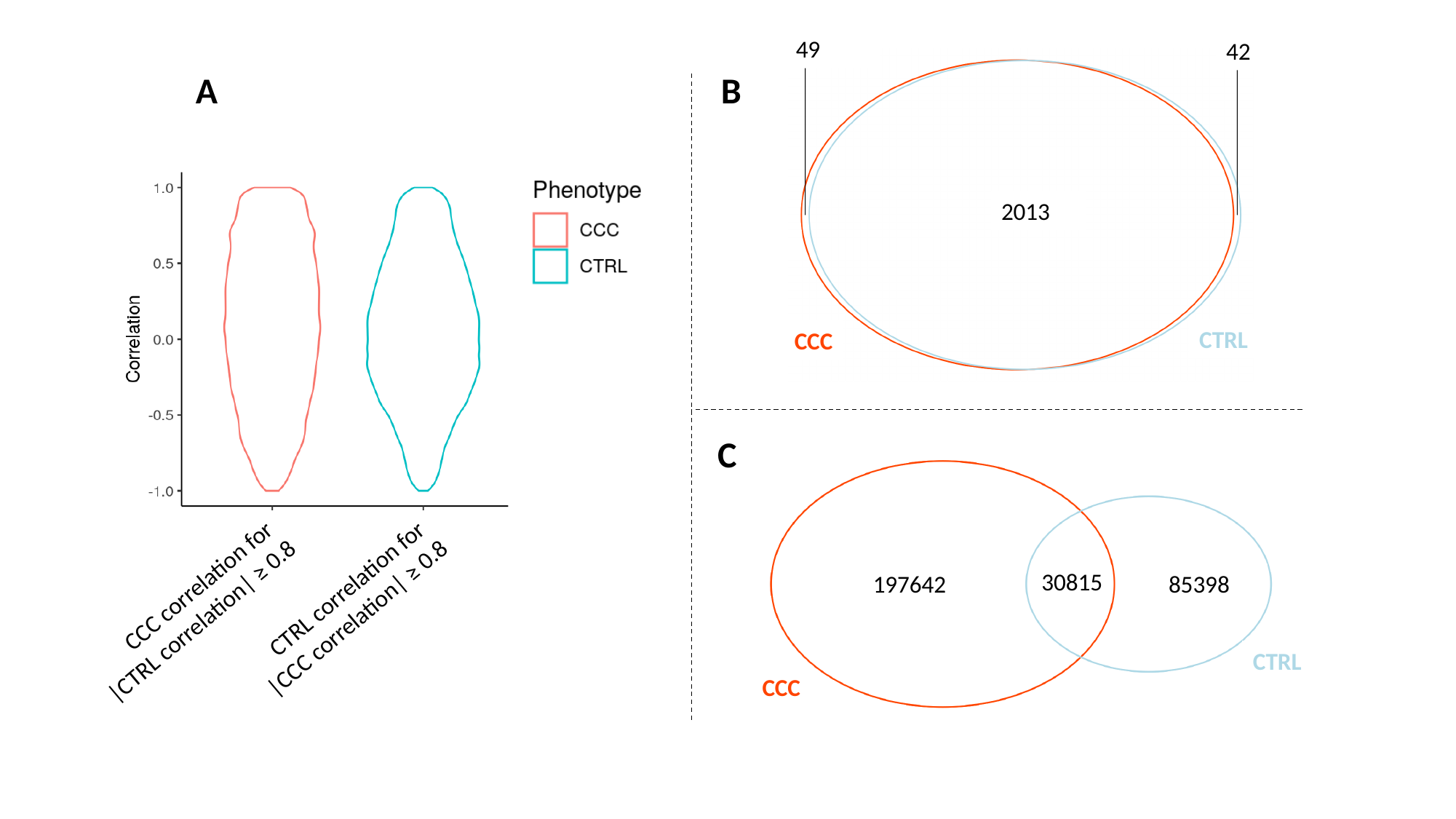

49
42
A
B
2013
CTRL
CCC
C
30815
197642
85398
CTRL correlation for
|CCC correlation| ≥ 0.8
CCC correlation for
|CTRL correlation| ≥ 0.8
CTRL
CCC
