## Supplementary figures and images for "Exploring global and specific pathogenic mechanisms in Chronic Chagas Cardiomyopathy through multi-omics integration"

### Supplementary figure 1.pptx

#### Slide 1
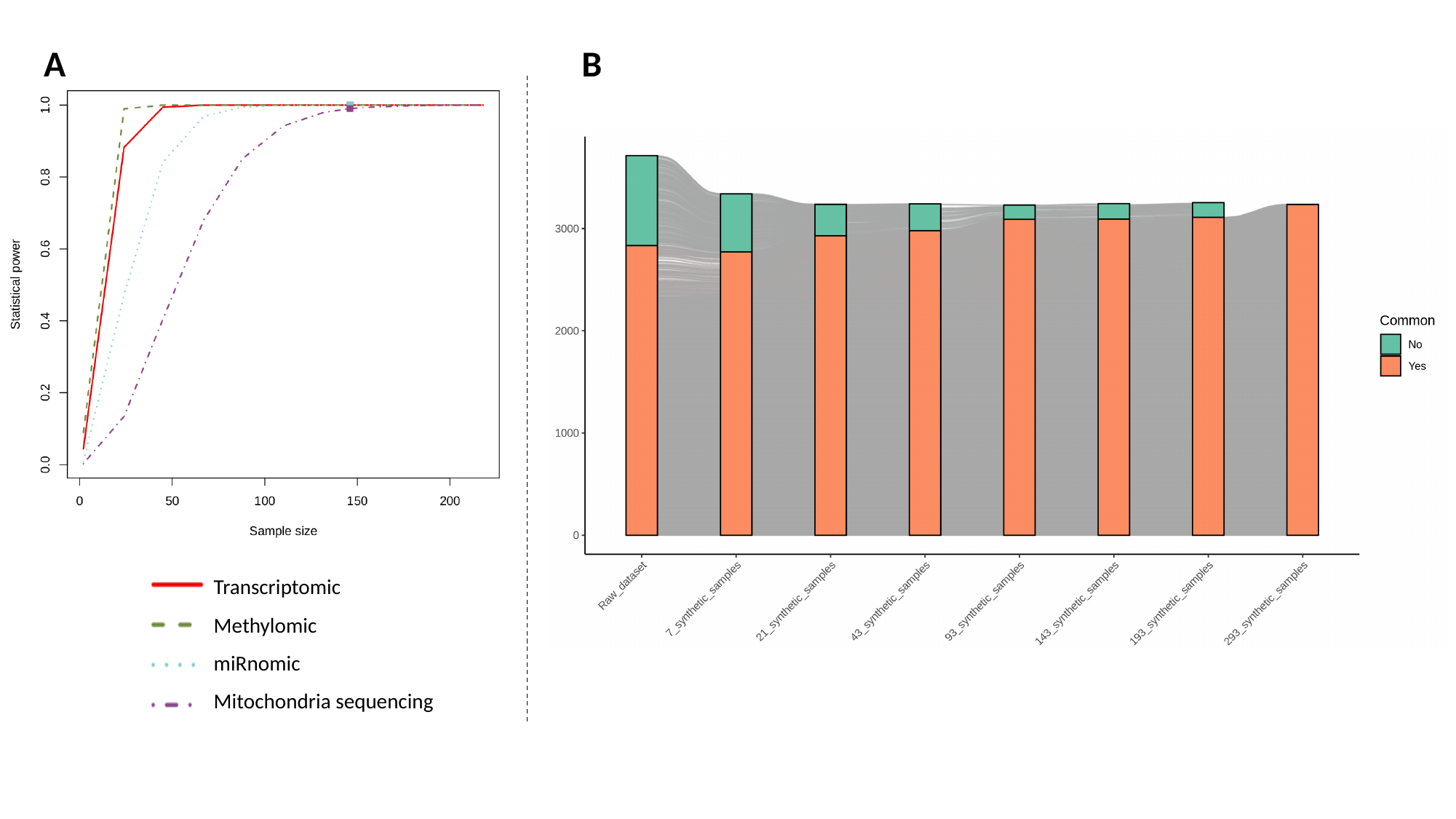

A
B
Transcriptomic
Methylomic
miRnomic
Mitochondria sequencing

### Supplementary figure 2.pptx

#### Slide 1
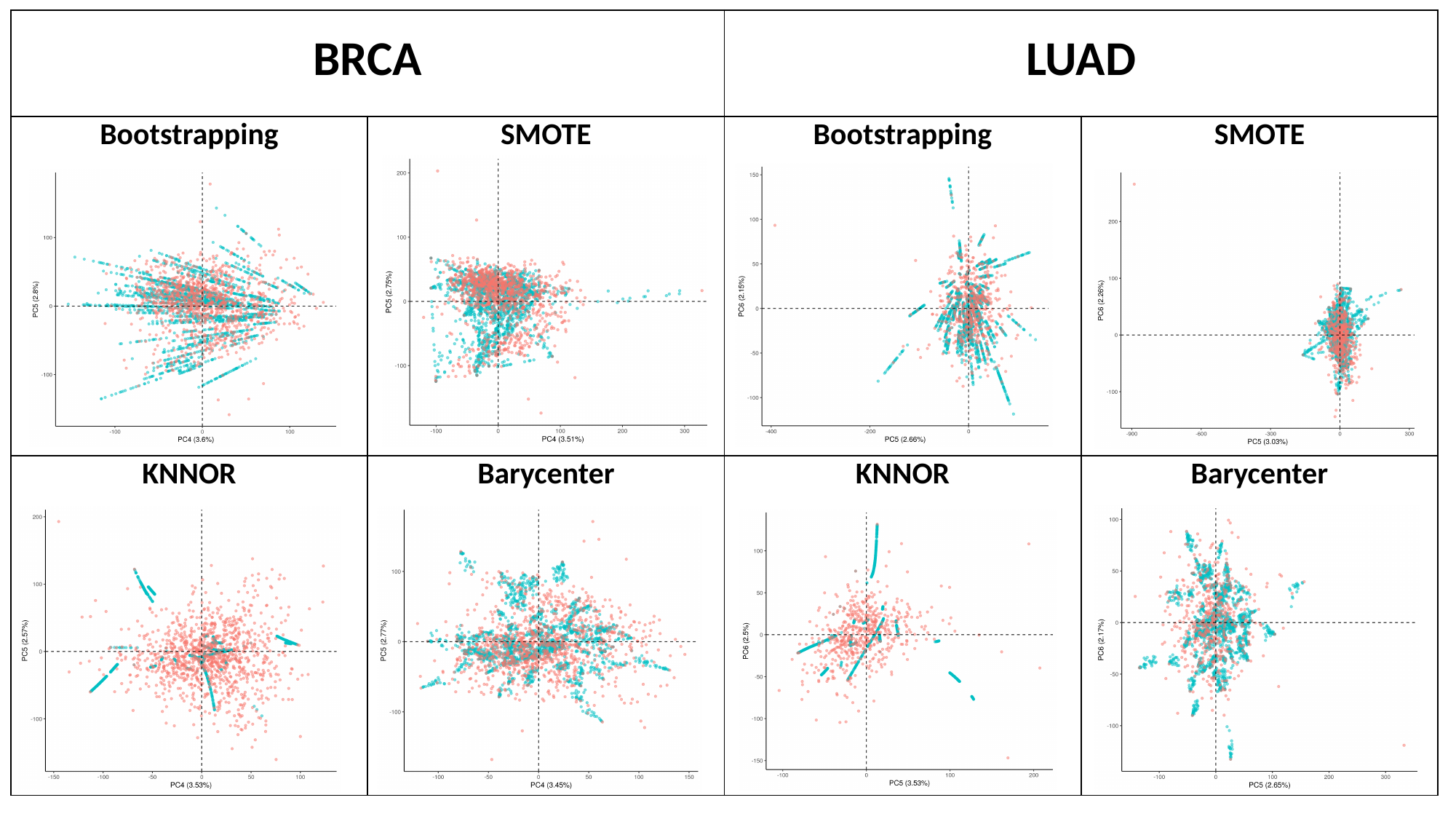

| BRCA | | LUAD | |
| --- | --- | --- | --- |
| Bootstrapping | SMOTE | Bootstrapping | SMOTE |
| KNNOR | Barycenter | KNNOR | Barycenter |

### Supplementary figure 3.pptx

#### Slide 1
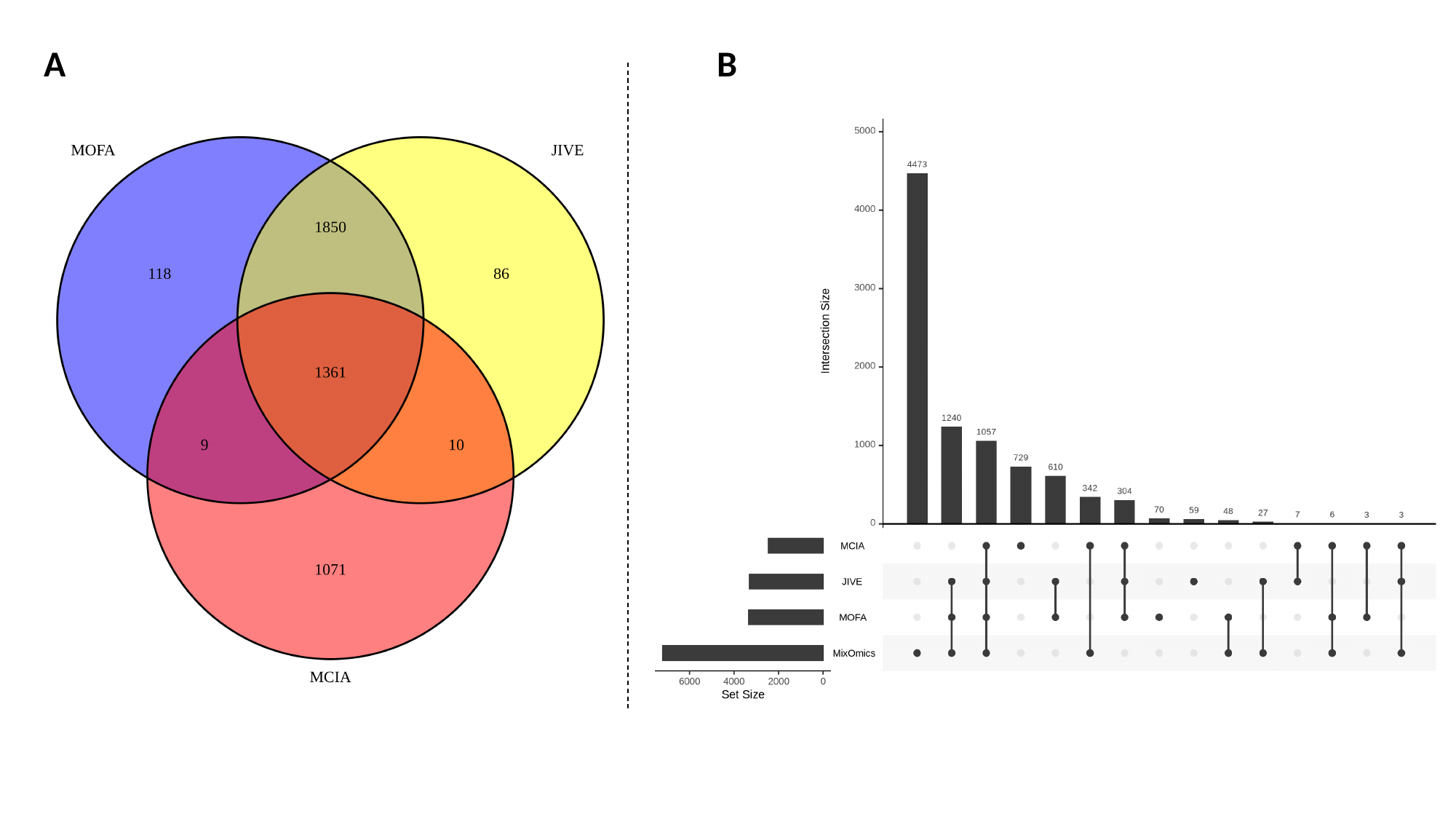

A
B

### Supplementary figure 8.pptx

#### Slide 1
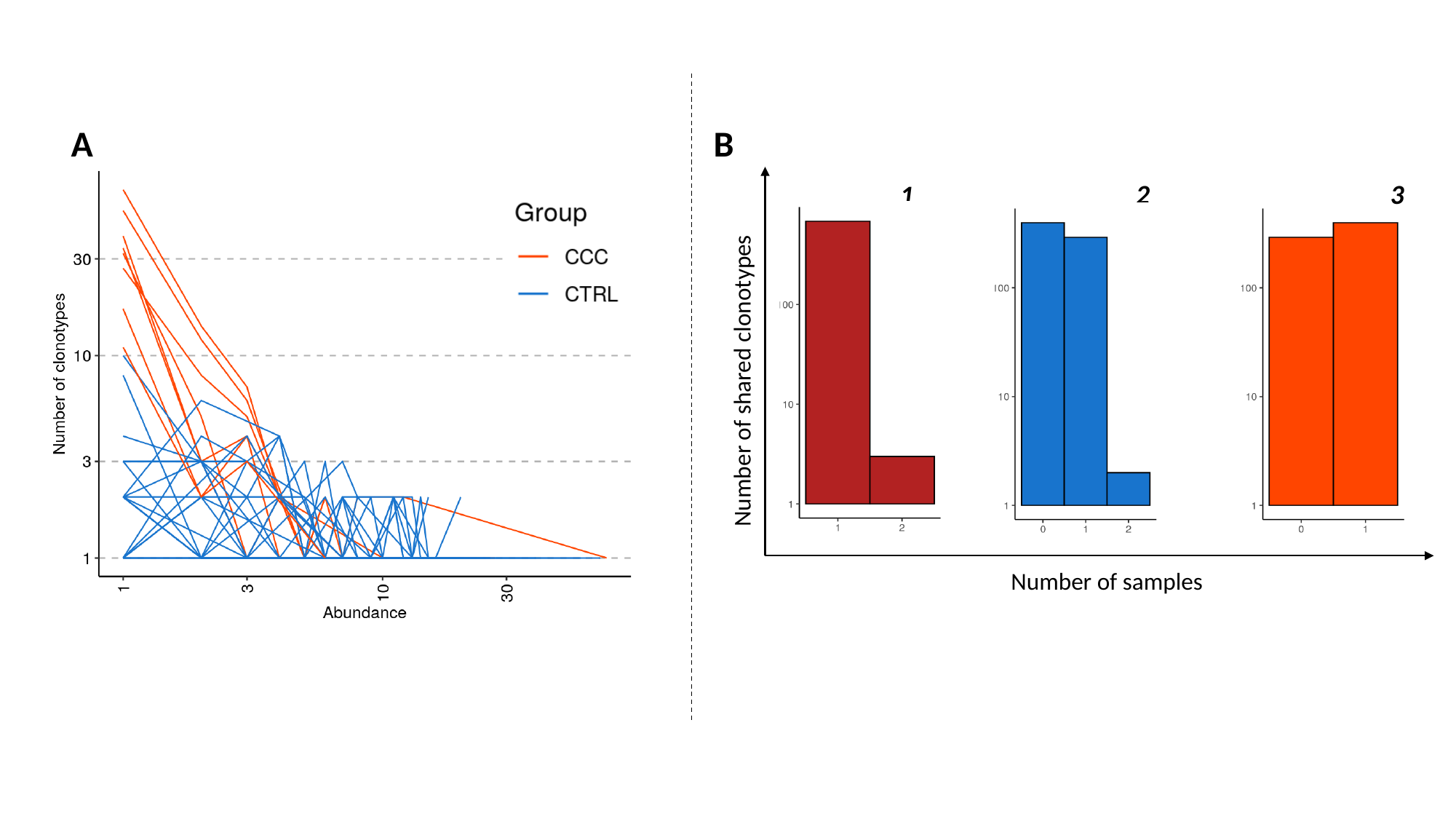

A
B
2
3
1
Number of shared clonotypes
4
Number of samples

### Supplementary figure 9.pptx

#### Slide 1
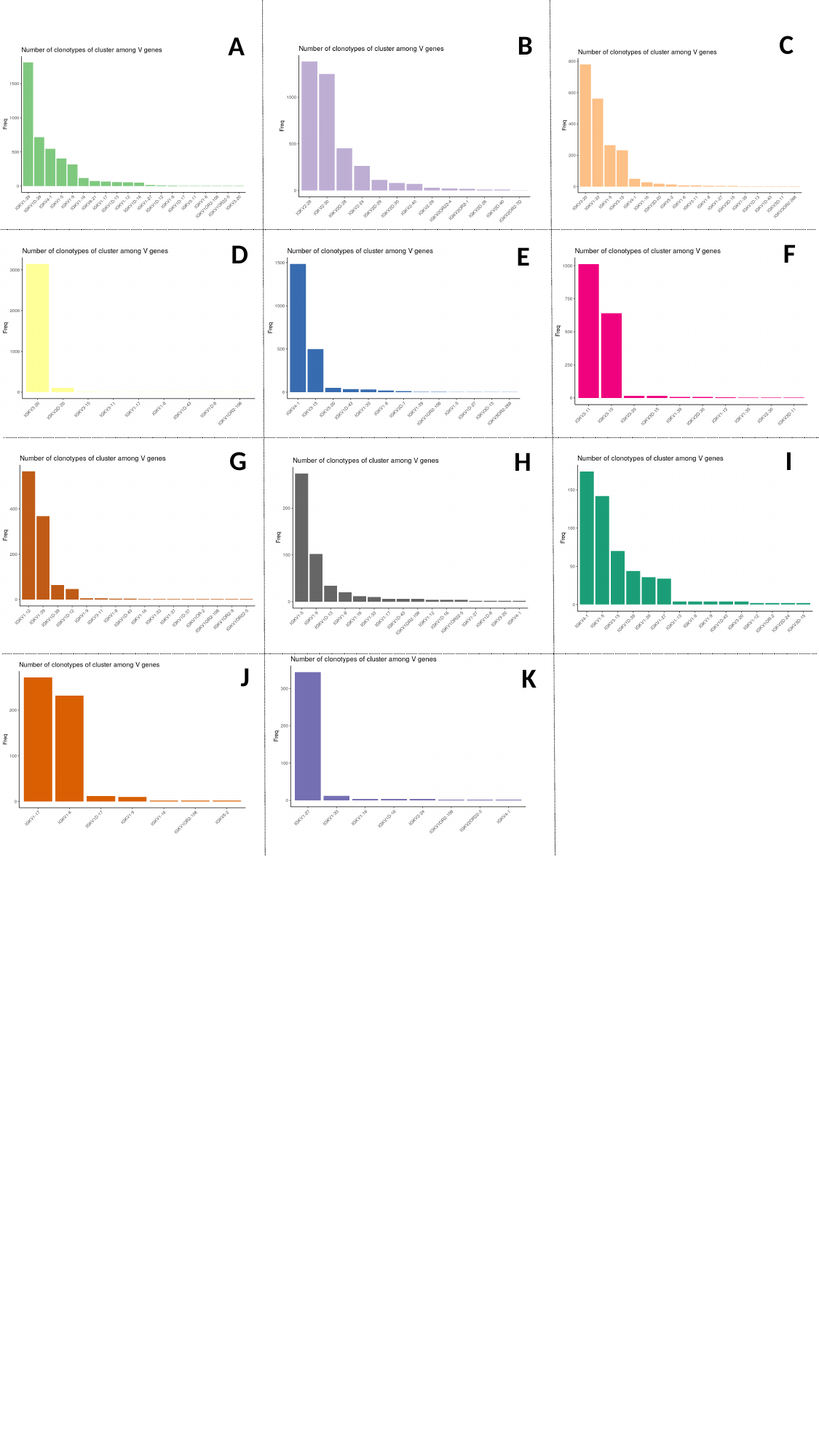

C
B
A
F
D
E
G
I
H
J
K

### Supplementary figure 10.pptx

#### Slide 1
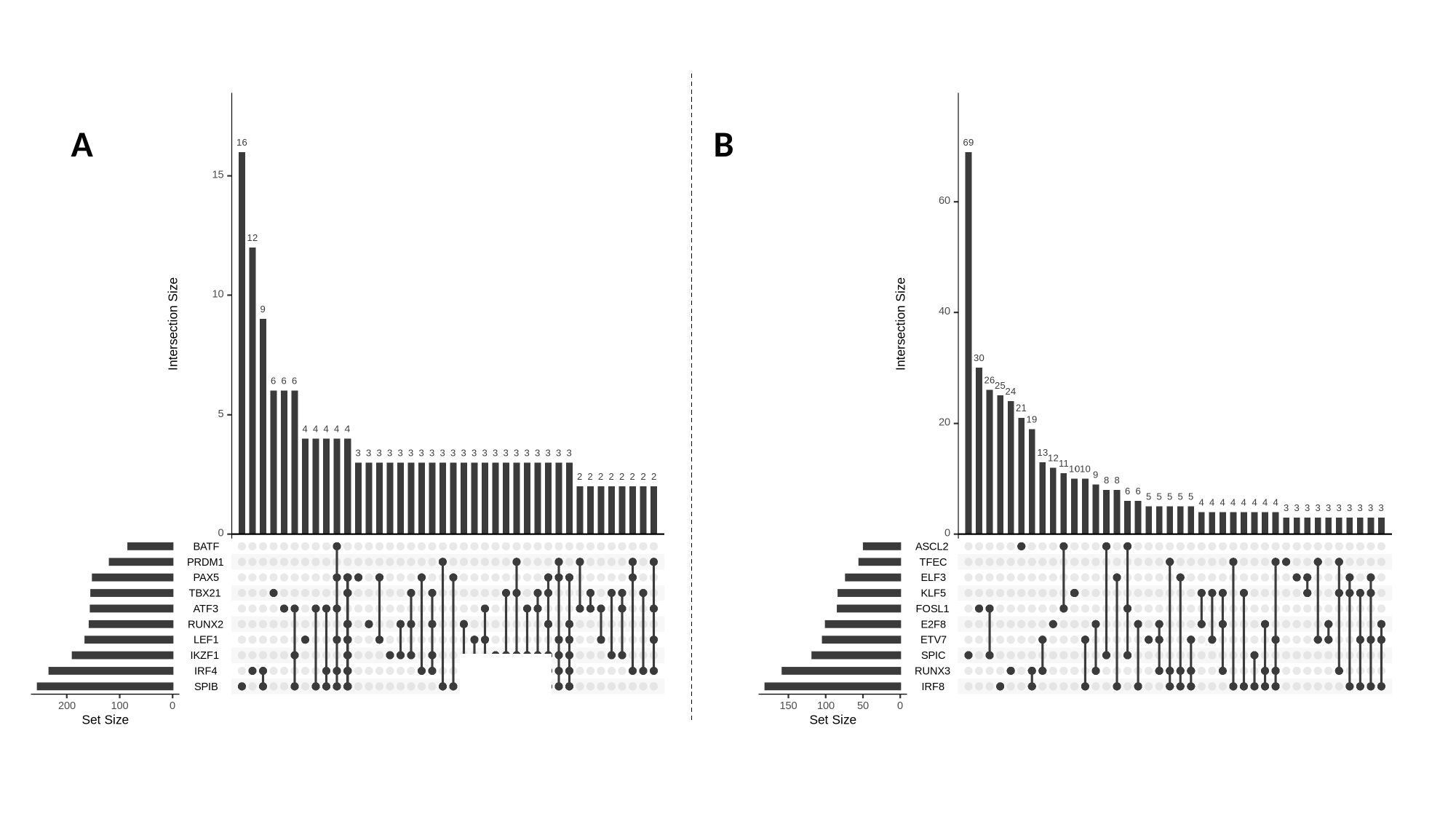

A
B

### Supplementary figure 11.pptx

#### Slide 1
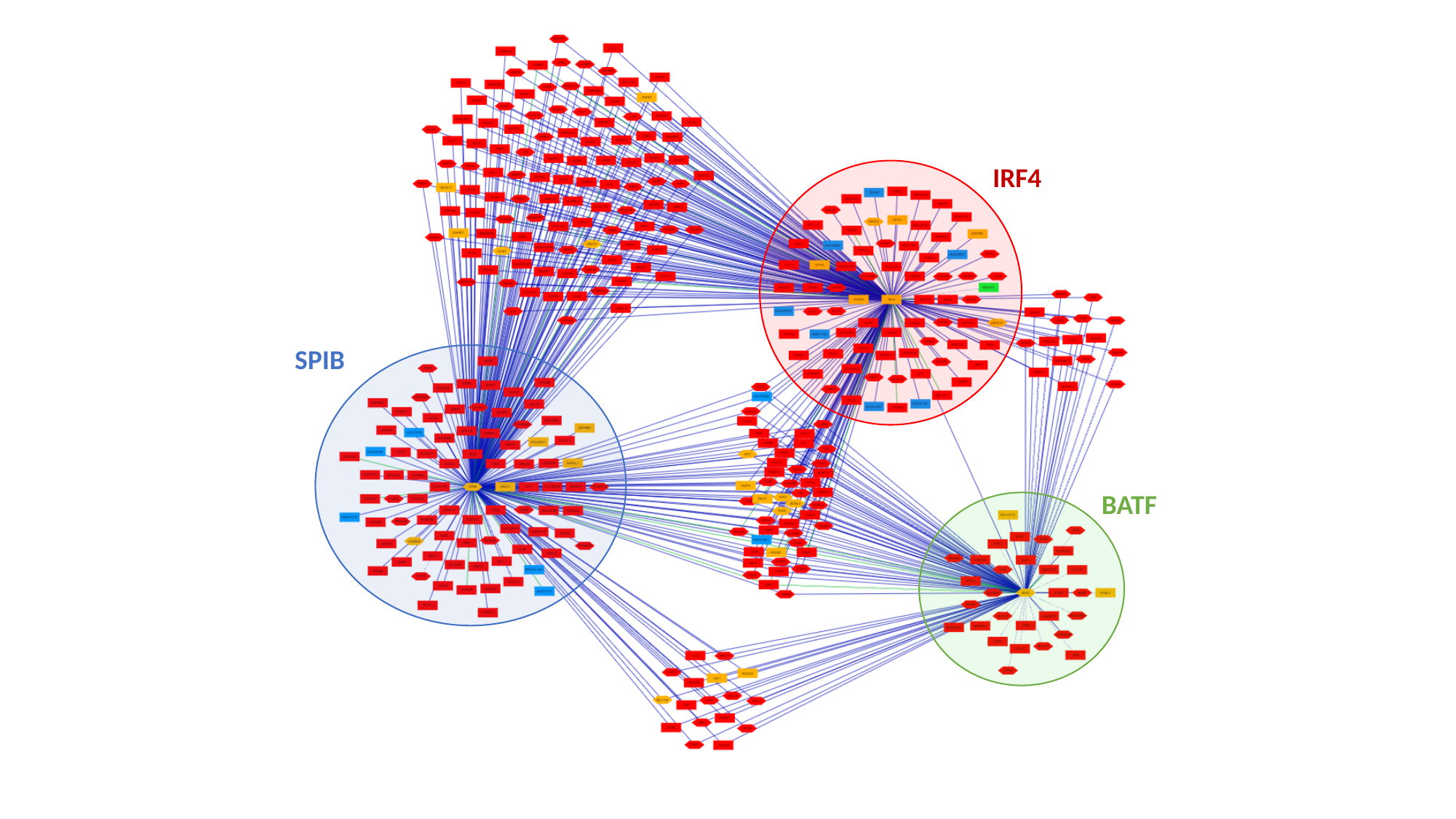

IRF4
SPIB
BATF

### Supplementary figure 12.pptx

#### Slide 1
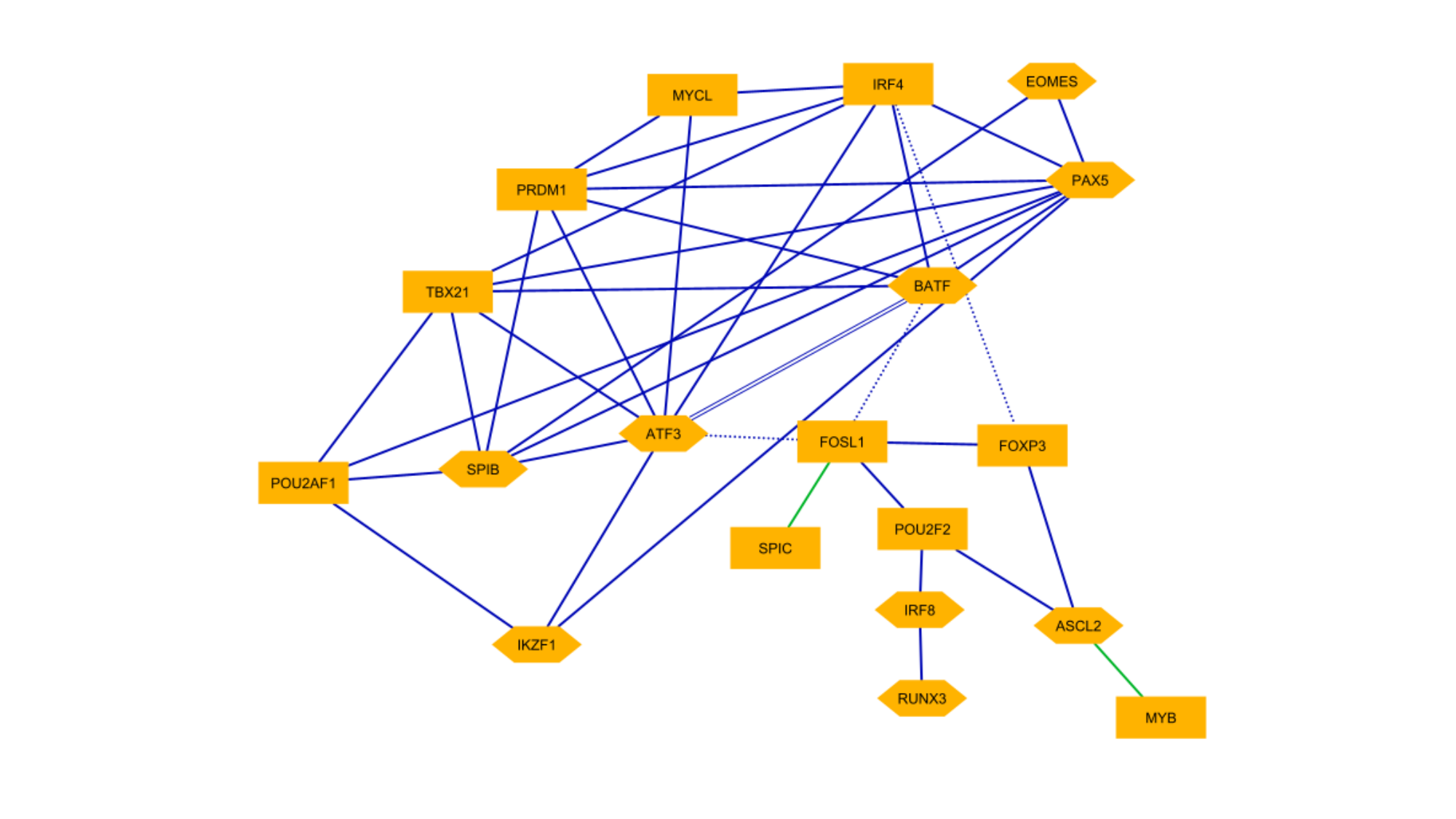

### Supplementary figure 13.pptx

#### Slide 1
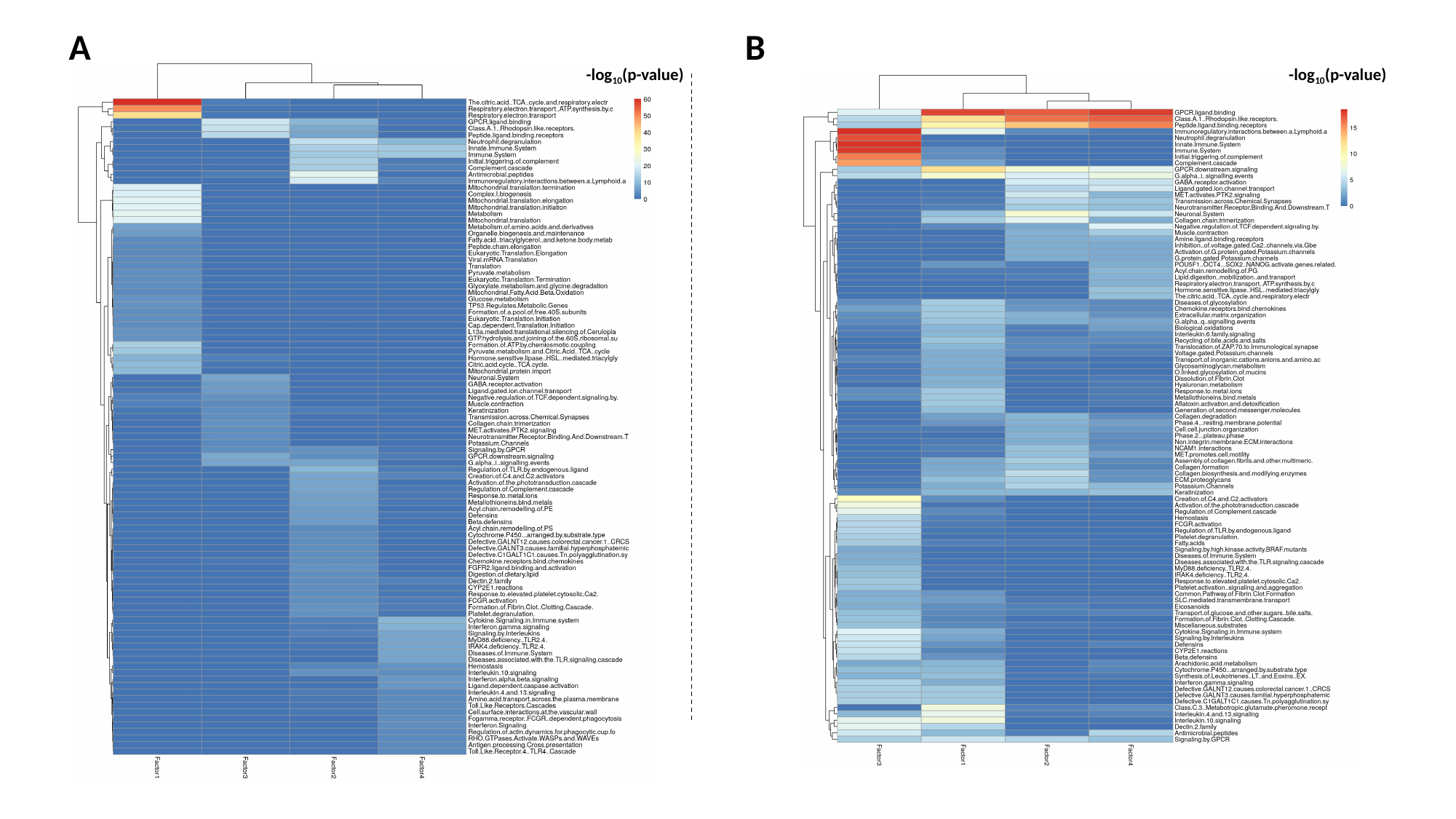

B
A
-log_10_(p-value)
-log_10_(p-value)
